# Risk assessment for long and short range airborne transmission of SARS-CoV-2, indoors and outdoors, using carbon dioxide measurements

**DOI:** 10.1101/2021.05.04.21256352

**Authors:** Florian Poydenot, Ismael Abdourahamane, Elsa Caplain, Samuel Der, Jacques Haiech, Antoine Jallon, Inés Khoutami, Amir Loucif, Emil Marinov, Bruno Andreotti

## Abstract

A quantitative analysis of the viral transmission risk in public spaces allows us to identify the dominant mechanisms that a proactive public health policy can act upon to reduce risk, and to evaluate the reduction of risk that can be obtained. The contribution of public spaces to the propagation of SARS-CoV-2 can be reduced to a level necessary for a declining epidemic, i.e. an overall reproduction rate below one. Here, we revisit the quantitative assessment of indoor and outdoor transmission risk. We show that the long-range aerosol transmission is controlled by the flow rate of fresh air and by the mask filtering quality, and is quantitatively related to the CO_2_ concentration, regardless the room volume and the number of people. The short-range airborne transmission is investigated experimentally using dedicated dispersion experiments performed in two shopping malls. Exhaled aerosols are dispersed by turbulent draughts in a cone, leading to a concentration inversely proportional to the squared distance and to the flow velocity. We show that the average infection dose, called the viral quantum, can be determined from epidemiological data in a manner consistent with biological experimental data. The results provide quantitative guidance useful for making rational public health policy decisions to prevent the dominant routes of viral transmission through reinforced ventilation, air purification, mechanical dispersion using fans, and incentivizing the wearing of correctly fitted, quality facial masks (surgical masks, possibly covered by another fabric mask, or non-medical FFP2 masks). Taken together, such measures significantly reduce the airborne transmission risk of SARS-CoV-2.

## 1 Introduction

Respiratory pathogens are transmitted via droplets emitted by coughing or sneezing. However, oral fluid droplets harbouring pathogenic particles are also generated during expiratory human activities (including breathing, speaking or laughing), which may cause asymptomatic and pre-symptomatic transmission. The atomization process producing aerosols occurs in the respiratory tract when an air flow of sufficient velocity leads to the fragmentation of a mucus film. Pathogens responsible for illnesses such as influenza, tuberculosis, measles or SARS can be carried by these small droplets, which can remain airborne for long periods of time [1–3].

There is ample evidence that SARS-CoV-2, the virus causing coronavirus disease 2019 (COVID-19), can be spread through aerosols [4–7]. Airborne particles are the dominant transmission pathway of COVID-19 in public places where respiratory mask wearing is compulsory. Indeed, the heavier, millimetre-sized droplets, known as sputters in common parlance, have a ballistic trajectory that is relatively insensitive to the presence of air and are stopped by all types of masks. Transmission through contact with fomites on which these drops have been deposited is probably not significant [8–10] – regardless of its actual weight in SARS-CoV-2 transmission, the improvement of hand hygiene remains a recommended habit to prevent the transmission of other pathogens. Finally, the possible transmission by faeces via aerosolization when toilets are flushed remains controversial and probably an ancillary pathway, if relevant at all. Airborne particles are dragged along by the motion of air; when turbulent fluctuations are able to keep the droplets in the air, they are known as aerosols. Water in the droplets then evaporates into the air, concentrating the droplets in virions and proteins from mucus, some of which have antiviral properties that help inactivate the virus after a few hours.

Viral particles have been directly evidenced in the air exhaled by patients, which can survive for several hours in a mucus droplet and remain airborne. SARS-CoV-2 has even been found in hospital COVID ward ventilation exhaust filters [11]. Animal model experiments have shown that SARS-CoV-2 can spread through the air in conditions where ballistic drops are excluded [12]. Transmission in the most detailed case studies [13, 14] can only be adequately explained through airborne spread. Long distance transmission in quarantine hotels has been documented [15], where the absence of close contacts was established via video surveillance footage review and the contamination chain was supported by genomic evidence. Asymptomatic, infected individuals do not cough or sneeze, yet they account for at least 50 % of all transmissions [16]; this suggests that they do not spread the disease via large ballistic droplets. Indoor transmission is 19 times more prevalent than outdoors [17]. The trajectories of large ballistic droplets are insensitive to indoor versus outdoor environments, whereas that of aerosols are not. The lack of long-range airborne infection outdoors provides a direct explanation for the risk difference indoors and outdoors. Moreover, good ventilation was shown to decrease transmission [5]. Healthcare workers wearing personal protective equipment designed to protect against ballistic droplets, but not aerosols, have been infected [18]. Finally, superspreading events, when a single patient infects a large number of people [19] can only be explained by a long-range transmission. All these arguments provide evidence for the airborne transmission of SARS-CoV-2.

Here, we revisit the problem of measuring the viral transmission risk [20] in public places such as schools, offices, university lecture halls, museums, theaters or shopping centers, but also outdoors. Our aim is to characterize the dominant transmission routes in social activities and to identify efficient ways of reducing the risk of epidemic contamination in public spaces. We first define the risk of transmission in a public space and document its dependence on the number of people present, the average time they are present, the available volume in which aerosols are stored and the level of ventilation.

We then discuss what an acceptable residual risk might be and against which ethical standard it can be assessed. The risk assessment for long-range airborne transmission, which is specific to indoor conditions and inexistant outdoors, is determined and quantitatively related to the CO_2_ concentration. We then show that short-range airborne transmission, localised in the wake of people infected by COVID-19, obeys the same physical laws indoor, and outdoor. We report experimental measurements of turbulent dispersion of a passive tracer (CO_2_) performed in two French shopping centers: Forum des Halles in Paris and Carré-Sénart. In most such public spaces, the turbulent diffusion is due to a small permanent air flow leading to a rapid spatial decay of the tracer concentration. The supplementary risk when staying in the wake from other people is determined quantitatively as a function of the distance downwind. From this risk assessment, we define quantitative standards (occupation capacity, CO_2_ level, ventilation, masks) which should be implemented in public spaces to reach the acceptable residual risk. In conclusion, we elaborate on various techniques available to reduce the viral transmission risk in public places, as a complement to vaccination.

## 2 An overview of the biology of SARS-CoV-2

We provide here an overview of the biology of SARS-CoV-2 for non-specialists. Although it provides the basis for risk assessment calculation, this section is mostly independent of the rest of the article and can be read afterwards as well.

### 2.1 Viral infection mechanism

SARS-CoV-2 is a virus enveloped by a lipid bilayer in which the E, M and S proteins are inserted. The lipid membrane comes from the cell in which the virus replicated before being released, – but not from its plasma membrane. The virus contains one copy of the genomic viral RNA protected by a capsid, structured by the assembly of the nucleocapsid protein N. Viral particles measure 80 to 90 nm in diameter, and are decorated with an average number of 48 spike ptoteins (S) anchored in their envelope. The RNA genome encodes 26 proteins, including the envelope (E, M and S) and capsid (N) proteins, as well as non-structural proteins that are necessary for the replication and the assembly of the virus inside the host cell [21, 22]. To colonize a cell, the virus, interacts through the S protein – which is cleaved by a host cell protease (the TMPRSS2 protease) – with a host cell membrane protein (the ACE2 receptor). Cleavage of the S protein is necessary for a conformational change so that it can effectively interact with the ACE2 receptor. This interaction leads to the formation of a virus-ACE2 complex which triggers the internalization of the virus inside the cell.

A series of cellular events lead to the disassembly of the virion and the undressing of the RNA molecule. The released viral RNA is taken in charge by the ribosomes of the host cell, which read the information it encodes and produce the twenty to thirty viral proteins needed to produce viruses. New viral particles are subsequently assembled, by hijacking the host cell mechanisms, and subsequently released, leading to the colonization of neighboring cells [23].

From the nasal cavity, which is probably the first tissue to be contaminated, the virus, embedded in the mucus secreted by certain cells of the nasal epithelium, is carried to the throat, then to the trachea and finally to the lungs or the esophagus, and finally to deeper organs. The severity and variety of symptoms of the disease depends on the likelihood that viral infection overcomes host defenses and reaches multiples sites, as well as on host-inflicted damages by the potent inflammatory and interferon responses launched against viral assault. In contrast, dissemination of the virus depends essentially on its ability to colonize the host respiratory tracts, and may thus not be correlated with the severity of symptoms [24]. From the nasal cavities, the virus has the possibility to go up to the brain and to colonize certain cells of the cortex. More rarely, the virus can be found in the blood or lymph, reach the different organs of the body and colonize specific cell types of certain organs (liver, kidney, heart, prostate, etc.) [25].

When the virus is concentrated in the nasal cavity or throat, it is disseminated via a mist of fine droplets of mucus or saliva dispersed by breathing, talking or singing. A sneeze or cough produces larger droplets containing viral particles. When these droplets are formed, their content in viral particles increases linearly with the viral charge in the nasal cavity for mucus droplets or in the throat for saliva droplets [26]. An organism may become infected if a sufficient amount of viral particles interact with cells expressing both the TMPRSS2 protease and the ACE2 receptor and if the virus is able to hack into cellular mechanisms to produce and disseminate new virions. At each cycle, the virus needs to enter cell, replicate its RNA molecule, produce the proteins required for its sefl-assembly, and then be released.

Omics database provide an overview of the tissues and cells types that express the TMPRSS2 protease and the ACE2 receptor in the different tissues of the human body (Human Cell Atlas). The commonly accepted hypothesis is that these cells constitute the first target cells for the SARS-CoV-2 virus [27]. The existence of a set of nasal cells in the expression map of the ACE2/TMPRSS2 suggests that the nasal epithelium is most often the first contaminated tissue [28–30].

### 2.2 Antiviral responses

The arrival of the virus in the nasal cavity induces an antiviral response. This response is due to the recognition of pieces of the virus by cells of the organism and involves the interaction of pieces of RNA or proteins of the virus considered as danger signals with cell receptors interacting with these danger signals. This interaction triggers a non-specific antiviral defensive response, which relies notably on the production of a class of molecules called interferons. Upon binding of their receptors on cognate target cells, interferons trigger the expression of a broad array of interferon-stimulated genes (ISGs) which, through a variety of mechanisms, contribute to restricting viral replication [31].

Interferons are produced by infected cells but also by sentinel immune cells. After their secretion, they diffuse and bind their receptors on surrounding cells without discriminating whether they are infected or not, shutting down cell functions. As concentration is higher around the site of infection, diffusion leads to an efficient stochastic tracking of infected cells. If the interferon response is launched early, in a localized and circumscribed way, the viral spreading across the cell tissue can be stopped [32, 33]. However, an overly strong interferon response is not only antiviral but also destructive. Moreover, the viral-host crosstalk is further complexified due to the fact that some of the viral proteins counteract the host interferon responses and in a reciprocal way, the host interferon responses may amplify the virus infectivity [34].

### 2.3 Role of the mucus in the contamination

The nasal cavity is a battlefield where the replication of the virus and its inhibition are opposed in time and space. The cellular mechanisms allowing the replication and secretion of the virus as well as those leading to the interferon response are specific to each individual. Thus, the kinetics of the mechanisms allowing the replication and secretion of the virus characterizes the capacity of an individual to be more or less contaminator. Similarly, the kinetics of the interferon response correlates with an individual’s susceptibility to infection [35]. An important element often forgotten modulates these kinetics in the nasal cavity and participates in the immune response: the mucus. Mucus is composed mainly of water (95%), lipids and proteins (mucins) [36, 37]. It is secreted by specialized cells (goblet cells) that are also the cells infected by the virus. Mucus forms two layers over the nasal epithelium, a gel with a network of polymerized mucins anchored to the membrane of the epithelial cells and a solution that moves under the action of the cilia of the epithelial cells. The viscoelastic properties of mucus depend on the type of mucins [38] (there are 16 isoforms of mucins) and on the physicochemical environment (humidity, calcium concentration and pH). There is a synergy between the mucus and the immune system (mucosal immunity): the network created by the polymerized mucins traps particles down to sizes of a few hundred nanometers (virus size). Mucus also contains enzymes that can neutralize viruses and bacteria in a non-specific manner. Nasal microbiota, a community of bacteria inside the mucus, also plays a role in the defense against contamination by viruses or pathogenic bacteria. It must be emphasized that nasal mucus and saliva, which is also a specific mucus, have different compositions and physicochemical characteristics. Several problems remain open:

- The respective contributions of the mucus and interferon responses in the clearance of SARS-CoV-2 [39].
- The role of mucus in the formation of droplets and their contaminating character: concentration of the virus, size of the droplets, mixture of the mucus of two different individuals – mucus of the transmitter and mucus of the receiver [40].
- The evolution of the physico-chemical characteristics of the mucus as a function of age, temperature, humidity, pH, ionic concentration (in particular calcium which in high concentration condenses the mucin polymers) and pathology [41].

## 3 Defining and modeling the transmission risk

### 3.1 Contamination as a Poisson process

Contamination risk assessment requires the estimate of the probability of contamination under a given intake viral dose. The intake dose is the amount of viral particles inhaled by a person, cumulated over time. The simplest hypothesis is to assume an independent action of all inhaled viral particles, which means that a single virus can initiate the contamination. The probability that at least one virus particle manages to enter a cell and replicate is independent of the presence or not of others viral particles. However, more than one is statistically needed, as the probability that a single virus overwhelms the host immunity defences successfully is small, typically between 10^−6^ and 10^−5^. Contamination takes place when a single virion penetrates to a vulnerable locus where conditions are favorable. Although it remains possible that a cooperative attack of many virions is needed to evades host defenses to initiate infection, the independent action will be assumed in most of this paper. The intake viral dose *d* is defined as the amount of virus particles inhaled by a person. It increases with the time of exposure to the virus and with the concentration of viral particles in the air. For an individual having inhaled an intake dose *d*, the probability law of infection *p*(*d*) takes the form:

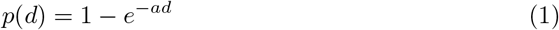

where the susceptibility *a* is the inverse of the infection dose defined, for each individual, as the intake dose for which the probability of infection is 1 − 1/e ≃ 63 %. This equation is characteristic of a Poisson process. *a* is widely distributed across individuals, according to a probability distribution *f* (*a*), for a given population. We denote by *ā* = ∫ *af* (*a*)d*a* the average of *a* over individual characteristics. *ā*^−1^ is called the quantum of infection for this population and can in principle be expressed as a number of viral particles, in GU (genom units). The dose *d* is standardly expressed directly in “quanta”, used as a convenient unit for a quantity of viral particles.

Cooperativity can be simply taken into account by assuming that *K* virions successfully overcoming the barriers are needed to replicate in cells and overwhelm the host antiviral defences. The probability of contamination then reads:

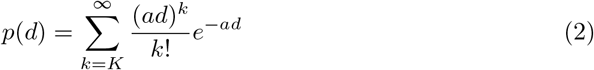

which resumes to the Poisson process for *K* = 1. As no evidence of cooperativity in viral infection has ever been provided, we will keep the simpler Poisson process hypothesis for the rest of the paper. It must not be confused with the Wells-Riley model, that will be discussed later on.

### 3.2 Viral load distribution

The viral load of an infected person depends on time. For simplicity, the concentration of viral particles in the exhaled air, noted 𝒟, can be assumed to present the same temporal profile amongst patients:

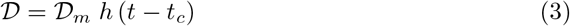

where 𝒟_*m*_ is a characteristic concentration and *t*_*c*_ the contamination time. Both 𝒟 and 𝒟_*m*_ can be expressed in GU*/*m^3^ or in quanta*/*m^3^. The rescaled viral load curve *h*(*t*) is dimensionless. *h* increases exponentially in the presymptomatic period and decreases exponentially at long time (Fig. 2). For reasons that will appear later, we choose here the following normalization for *h*(*t*), which provides an unambiguous definition of the mean contagious time *T* :

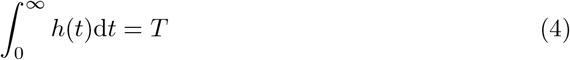

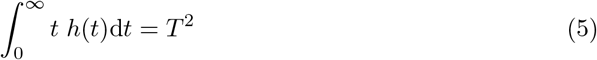

**Fig 1.**
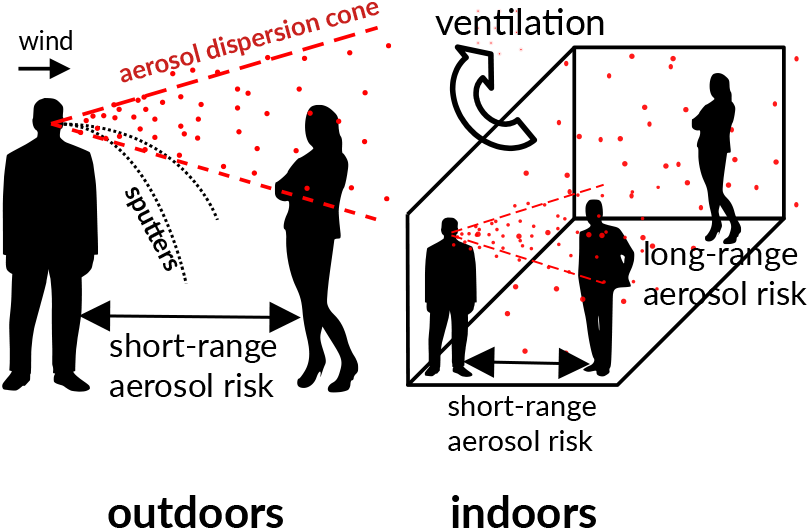
Graphical abstract. Outdoors, airborne viral transmission only takes place in the wake of an infected person: the exhaled breath is very concentrated in viral particles and is gradually dispersed by turbulent air fluctuations. The transmission risk typically decays as the inverse squared distance to the infected person. There is no long-range transmission oudoors. Indoors, the same turbulent dispersion induces a short-range risk but the finite volume leads to a finite dilution of viral particles, hence a long-range contamination risk.

**Fig 2.**
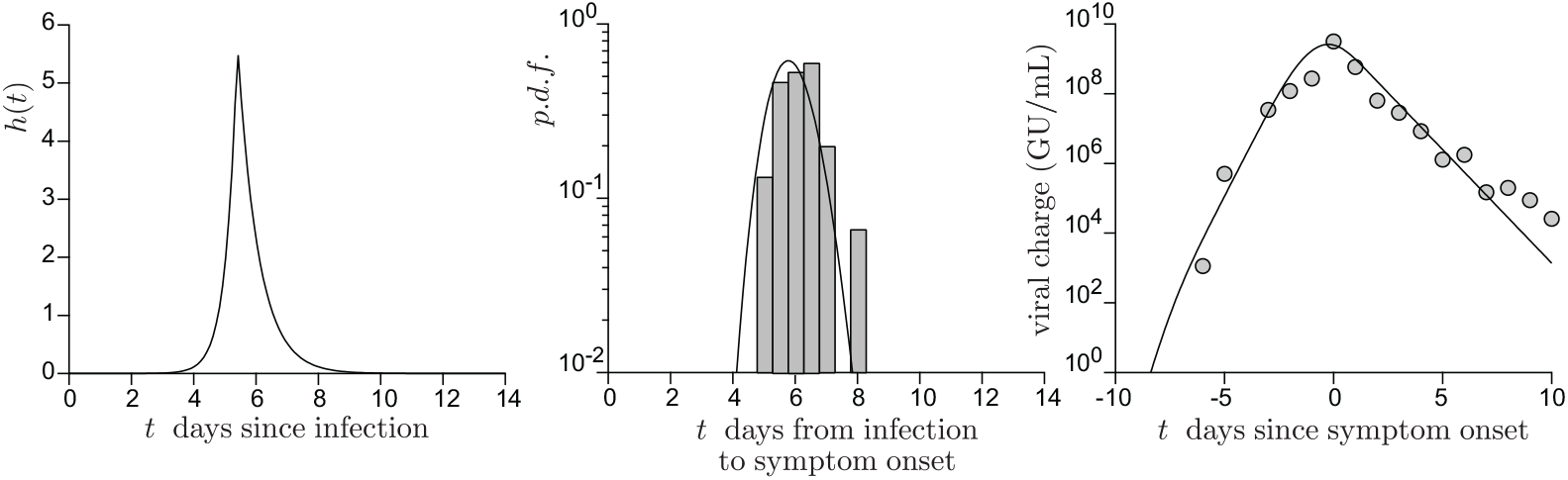
(a) Model dimensionless viral charge *h*(*t*) as a function of time *t*, in days, after contamination. *h*(*t*) is assumed to increase exponentially up to the maximal viral load and then to decay exponentially [42]. The best fit to the data provides the four fit parameters: the maximal viral charge is reached after 5.4 days; the exponential growth rate before this maximum is 2.8 days^−1^; the exponential decay rate after this maximum is −1.5 days^−1^. (b) Histogram of the duration between contamination and symptoms [43]. The solid line is the best fit by a Weibull distribution. (c) Average viral charge from Jang et al. [42] as a function of the duration after symptoms. The best fit provides the average viral charge 𝒟_*m*_.

The characteristic viral concentration 𝒟_*m*_ is highly variable amongst infected people. In order to consider an average over all statistical realizations, we introduce the probability density function *g*(𝒟_*m*_). As analyzed in Goyal et al. [44], the heterogeneity of 𝒟_*m*_ amongst people is related to the phenomenon of epidemic superspreading.

For the Wuhan-1 reference strain, the contagious time *T* is equal to 5.7 days with the above definition (Fig. 2). The maximum viral load is then *h*(0) 𝒟_*m*_ ≃ 5.6 𝒟_*m*_. The mean viral charge 𝒟_*m*_ is around 10^8^ GU for the wild strains [42, 45]. The mutant strain B.1.1.7 exhibit a double viral charge as compared to the wild-type (𝒟_*m*_ ≃ 2 10^8^) but its basic reproduction number is 1.5 larger only. This increases the viral persistence in the sense that a longer duration (+2.2 days) above the chosen threshold for the RT-PCR tests is observed. However, the contagious time *T* does not increase. The mutant strains P.1 and B.1.617 present a basic reproduction number 2 times larger than the wild strains which, probably, is associated to a larger mean value of 𝒟_*m*_ [46].

### 3.3 The individual risk budget

From the point of view of a rational agent who computes their benefit-risk balance for any activity, risk would naturally be defined as the probability of being infected while attending a certain public space. This risk depends on the characteristics of their immune system, on their co-morbidity factors and on the average viral dose they statistically inhale in this public space. For such a (non)rational agent, the risk is proportional to the epidemic prevalence *P* defined as the fraction of infected people. This type of individualist reasoning constitutes the central reason for which the epidemics presents stop-and-go waves.

A truly rational agent must therefore be altruist and base their benefit-risk balance on the number of people they would contaminate if asymptomatic or presymptomatic. In order to sustain an epidemic decay (Zero Covid strategy), each COVID-19 infected person must contaminate less than one person on average during the time they are contagious, hence in the range of 7 days for the wild strain and 10 days for the B.1.1.7 and P.1 mutant strain. A crude proportionality relation provides an estimate of the maximum acceptable risk per unit time around 0.0035 hour^−1^. This must be understood as follows: if during each hour, the risk to contaminate someone, being asymptomatic or pre-symptomatic, is limited to 0.35 % then the contribution to the creation of chains of contamination remains consistent with an epidemic control. In other words, amongst 300 infected people attending a certain public space during one hour, only one would be statistically allowed to contaminate someone else. The calculation can be refined by separating private, intra-family time, without masks, and the fraction of time spent in public space, wearing a mask. Intra-family contamination can be fought by a combination of testing, isolation and ventilation; however, it remains overall more effective than contamination in public space. A crude approximation would be to consider that any extra-family contamination leads to the contamination of the whole family, multiplying, in France, the contaminations by a factor 2.2 – the average size of a family. Estimating the average public time around 80 hours in 10 days, the acceptable risk per unit time in a public space becomes 0.005 hour^−1^: one infected person over 200 must, at maximum, infect another person during one hour.

This idea of an individual risk budget, even considering an altruist rational agent, belongs to the hygienist, behaviorist tradition. A scientific definition of the risk should include the partial environmental and social determination of behaviours.

### 3.4 A definition of the transmission risk in a public space

From a scientific perspective, the risk induced by a certain public space should characterize its contribution to the epidemic reproduction rate *R*, defined as the average number of people infected by a virus carrier. This risk should not depend on the epidemic incidence rate but only on the statistical creation of chains of contamination inside this public space, given the health procedures. It is, indeed, the epidemic reproduction rate that determines whether the epidemic amplifies (*R >* 1) or decays (*R <* 1). Here, we therefore define the risk of a public space as the average number of infections *r* that a COVID-19 infected person would cause on the average by staying in it. In order to be useful, this risk *r* must be directly comparable to 1. The individual risk, defined for an altruist rational agent, would then be the average of the risk *r* over a time window corresponding to the infectious time, weighted by the duration of each social activity.

Considering the contamination as a Poisson process, the mean number *Z* of people contaminated in a certain public place hosting *N* people during a given period of time, amongst which *M* infected people reads:

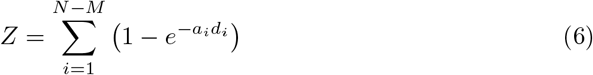

where *d*_*i*_ is the intake dose of the individual labelled *i* while 1*/a*_*i*_ is their contamination dose. In the case where all *N* people are statistically subjected to the same intake dose *d*_*i*_ = *d*, the risk *r* reads:

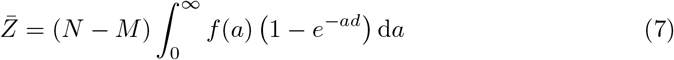

The quantity ∫*f* (*a*) (1 −*e*^−*ad*^ ) d*a* is the probability to get infected when an intake dose *d* is inhaled. It is called the dose response function. We consider now the low limit where the intake dose, expressed in quanta *ād* has a very low probability of being larger than 1. This excludes super-spreading events, which occur when an infected person with a large exhaled concentration 𝒟_*m*_ attends an under-ventilated place, leading to multiple simultaneous infections. Then, performing the linearization 1 − exp(−*ad*) ≃ *ad*, the equation simplifies to:

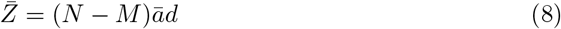

The average number of secondary infections is proportional to the intake dose expressed in quanta, *ād*, and to the number of non-immune people (*N* −*M*).

We now introduce the dimensionless dilution factor *ϵ* between the viral concentration in the inhaled air and the viral concentration in exhaled air 𝒟. In the next section, we will discuss how *ϵ* can be related to CO_2_ concentration and to mask efficiency. To determine the risk, as defined above, the *M* infected people are statistically picked up amongst the *N* people present. The inhaled dose of one particular person amongst *N* can, averaged over configurations, be expressed as:

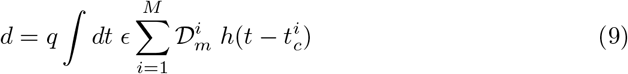

where *q* is the inhalation rate, i.e. the product of the breathing rate by the tidal volume. On the average, for light exercise, *q* ≃ 0.5 m^3^*/*hour.

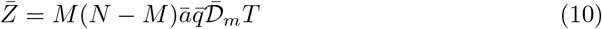

To evaluate the risk, the factor *M* (*N* − *M*) must itself be averaged over *M*. For simplicity’s sake, the fact that people from a same group are more likely to get infected together is ignored. The probability that *M* people are infected amongst *N* is governed by the binomial law, given the epidemic prevalence *P*. The mean number of infected people is 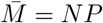. The mean number of infected people 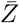 involves the multiplicative factor 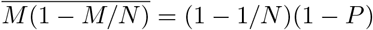. At small epidemic prevalence, the risk is independent of *P* as expected: it characterizes the creation of contamination chains.

The risk is proportional to the product of the mean inhalation rate 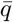 by the integrated viral concentration 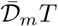 divided by the mean quantum of infection *ā*^−1^.

These four parameters can be combined into a single one, which is mean integrated quantum emission 𝒩 defined by:

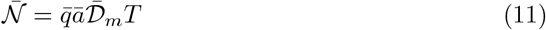

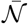 may depend on the particular activity taking place in the public space. The risk, as defined above, finally reads:

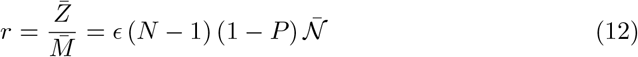

## 4 Long-range airborne transmission risk measured using carbon dioxide concentration

### 4.1 Relating the dilution ratio to carbon dioxide concentration

Consider first the case of a public space where nobody wears a respiratory mask. In first approximation, both carbon dioxide and aerosol droplets are emitted by an infected person at a rate proportional to the respiratory rate, which depends on their activity. When gravity is negligible in front of turbulence-induced drag forces, aerosol droplets are also dispersed in the air according to the same effective laws as CO_2_. We can safely consider that the *M* COVID-19-infected people are statistically picked up amongst the *N* people present, who all emit CO_2_. The dilution factor between the exhaled air and the air inhaled by one particular person amongst *N* can be expressed as:

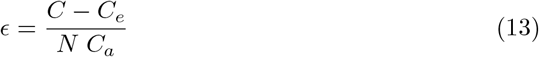

where *C*_*a*_ ≃ 37500 ppm is the average CO_2_ concentration in exhaled air.

Finally, we introduce a filtering factor *λ* due to the effect of respiratory masks, which will be quantified below. The use of air purifiers can be similarly included in the risk formula: just like masks, it does not change the CO_2_ level, but reduces the contamination risk.

We obtain the final formula for the average risk in a public space, in the low risk limit:

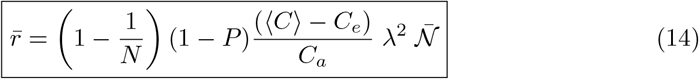

where ⟨*C*⟩ is the average concentration at the location where the (*N* −*M*) non-infected people stand statistically at time *t*. This equation, which constitute a central result of this paper, is remarkable: at large *N*, the dependence of the risk with respect to the volume of the room, the ventilation or the number of people are all encoded in the space averaged CO_2_ concentration ⟨*C*⟩.

The result is more subtle than it seems at first sight. Indeed, the *N* occupants of a public space all exhale CO_2_ but only the *M* infected ones exhale virions. Let us compare the risk of a well-ventilated lecture hall (say, at 750 ppm of CO_2_), with 50 students, to the risk if the same students are spread out in two conventional rooms with the same CO_2_ level. Obviously, all other things being equal, the probability of a student being infectious is twice lower when 25 students are grouped together, rather than 50.

However, since the ventilation must be proportional to the room occupation capacity to keep the CO_2_ level, the viral particles are twice more concentrated in the small room and therefore the inhaled doses double. Under the above assumption, the average number of people infected is the same, although the risk is distributed differently: when the intake dose *ad* is not small compared to 1, there is a larger probability to get an infection cluster (in the weak sense *Z >* 1) using two small rooms as the dose is higher.

More generally, the increase of the risk by a factor *N* due to the fact that *N* people can be infected is exactly balanced by the factor 1*/N* in the expression of the dose, related to the fact that not only infected people emit CO_2_, but everyone instead. For this, we need to linearize the equation with respect to the intake dose *ad*, an approximation which gets better and better as *N* increases and which constitutes an upper bound for the risk.

### 4.2 Long-range transmission: the well-mixed hypothesis

The long-range airborne transmission risk can be modeled using the well-mixed hypothesis: the viral particles are assumed to be dispersed over the whole enclosed volume *V*. Denoting by *Q* the flow rate of fresh air, the conservation of CO_2_ reads:

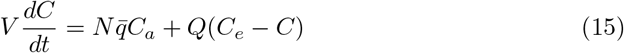

where *C* is the average CO_2_ concentration. Figure 3 shows a sample of the number of people *N*(*t*) in the Euralille shopping mall and the associated CO_2_ concentration signal *C*(*t*). The CO_2_ concentration is delayed with respect to number of people *N* so that ventilation is better controlled using *N* than *C*.

**Fig 3.**
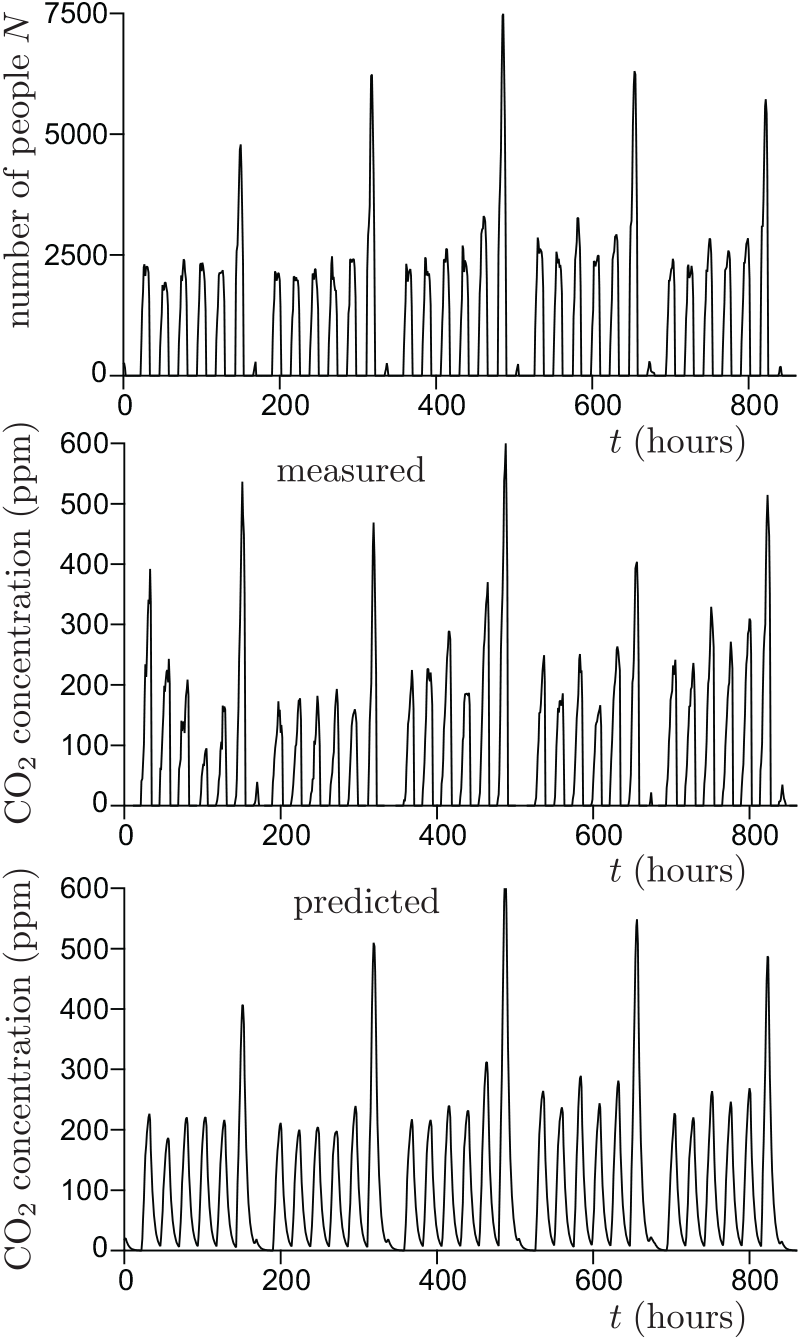
(a) Sample of the number of people *N*(*t*) in the EuraLille shopping mall in september 2020. (b) CO_2_ concentration *C* − *C*_*e*_ averaged over 10 sensors spread over the shopping mall. (c) CO_2_ concentration *C* − *C*_*e*_ predicted from equation (15) using the number of people *N*(*t*) as an input. The shopping mall volume and the total ventilation flow rate are measured independently.

Expressing *C* as a function of *ϵ* with equation (13), we get:

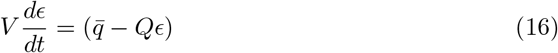

The sedimentation flux of heavy particles can be included inside *Q* as well. It is a linear relaxation equation towards the steady state solution:

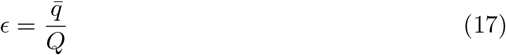

The exponential relaxation time is simply the ratio of the volume *V* to the fresh air flow rate *Q*.

### 4.3 Exponential epidemic growth

Consider a micro-society which shares the same air, under permanent social conditions. The epidemics will grow exponentially with a rate noted *σ*. Then the average risk is directly the epidemic reproduction rate *R*:

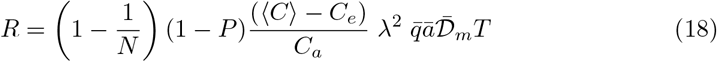

Contrarily to the previous formula, the transmission time is comparable to the epidemic timescale *σ*^−1^. The infection rate *I*, defined as the mean number of infected people per unit time obeys a Fredholm integral equation of second kind:

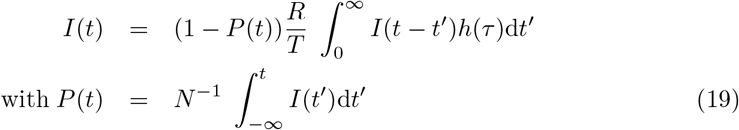

At small epidemic prevalence *P*, these equations admit an exact exponential solution. The growth rate *σ* is related to the epidemic reproduction rate *R* by the Lotka-Euler equation:

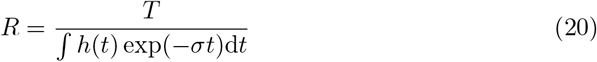

This relation is plotted in figure 4 for the rescaled viral load shown in figure 2.

**Fig 4.**
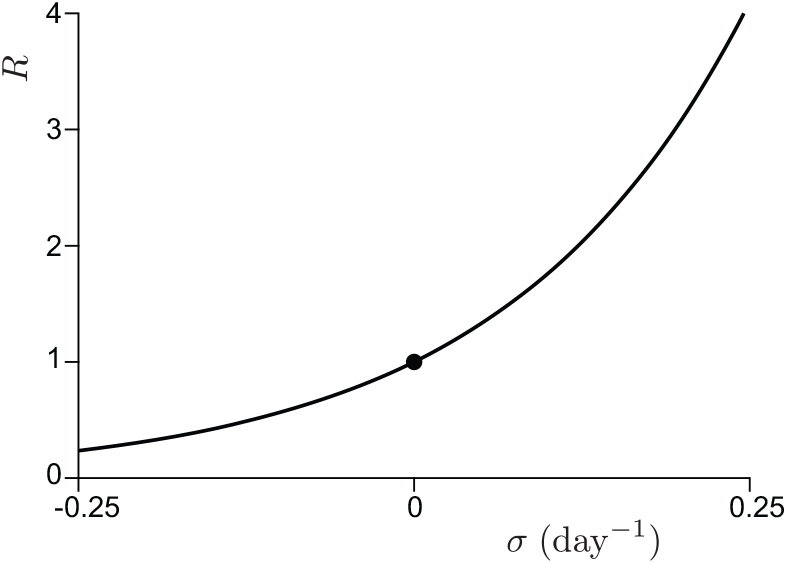
Relation between epidemic reproduction number *R* and the observed epidemic growth rate *σ*, as given by equation (20).

## 5 Short-range airborne transmission risk indoor and outdoor

The air carrying viral particles is gradually diluted by turbulent dispersion after exhalation by an infected person. In its vicinity, the concentration of viral particles is therefore higher than far away. In an indoor space, the dilution is limited by the finite volume, which leads to an accumulation of viral particles. Indoor and outdoor spaces differ by the long-range risk of transmission but share the short range one. Two transport mechanisms exist at short-ranges: ballistic droplets exit the nose and mouth with an initial momentum and rapidly fall to the ground under their own weight, while aerosol droplets are carried by an airflow, whichever is greater between the exit velocity out of the body and the ambient airflow. We investigate here the short-range aerosol risk by measuring and modeling the turbulent dispersion of CO_2_ in a generic situation. We make use of controlled experiments performed in different corridors of two French shopping malls, under various ventilation conditions.

### 5.1 Experimental set-up

The experimental setup is shown in figure 5. A controlled CO_2_ source of constant mass rate 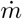 is obtained by sublimating dry ice sticks in an open cylindrical container of diameter 20 cm heated by a hot plate. The source is positioned at a distance 1.1 m above the ground. The measured sublimation rate 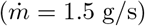 is equal to about 150 times the CO_2_ exhalation rate of an adult at rest. This allows us to measure CO_2_ concentrations with a high relative accuracy.

**Fig 5.**
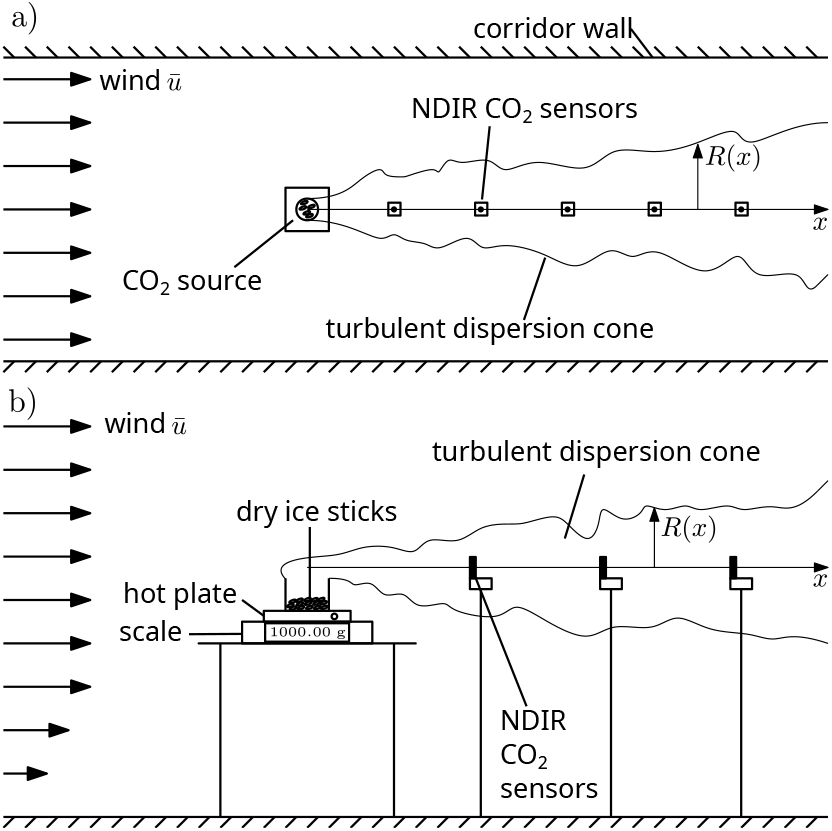
Schematic of the experimental setup used to measure the turbulent dispersion of contaminants. Dry ice sticks stored in an open cylindrical container are sublimated using a hot plate. The gas produced is convected downstream by the small draught blowing in the corridor where the CO_2_ concentration is recorded. The scales measures the mass injection rate 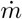. (a) Top view of the corridor layout. The source size (∼20 cm) is much smaller than the typical corridor width (5 to 10 m). (b) Zoomed-in view of the source. The sensors are placed at 1.1 m above ground.

The experiment had been designed, imagining that the turbulent dispersion would be statistically isotropic, as expected for an effective diffusion caused by large scale incoherent turbulent motion. Preliminary observations using a source of micron size glycerol+water droplets (Fig. 6), have shown that in most large public spaces, there are draughts causing horizontal transport and biased dispersion. After identification of the “wind” direction, non-dispersive infrared (NDIR) CO_2_ sensors are placed downstream from the source. They recorded the CO_2_ concentration *C* over the duration of each experiment, around 30 minutes. the initial and final values of *C* are used to determine the background CO_2_ level *C*_*e*_.

**Fig 6.**
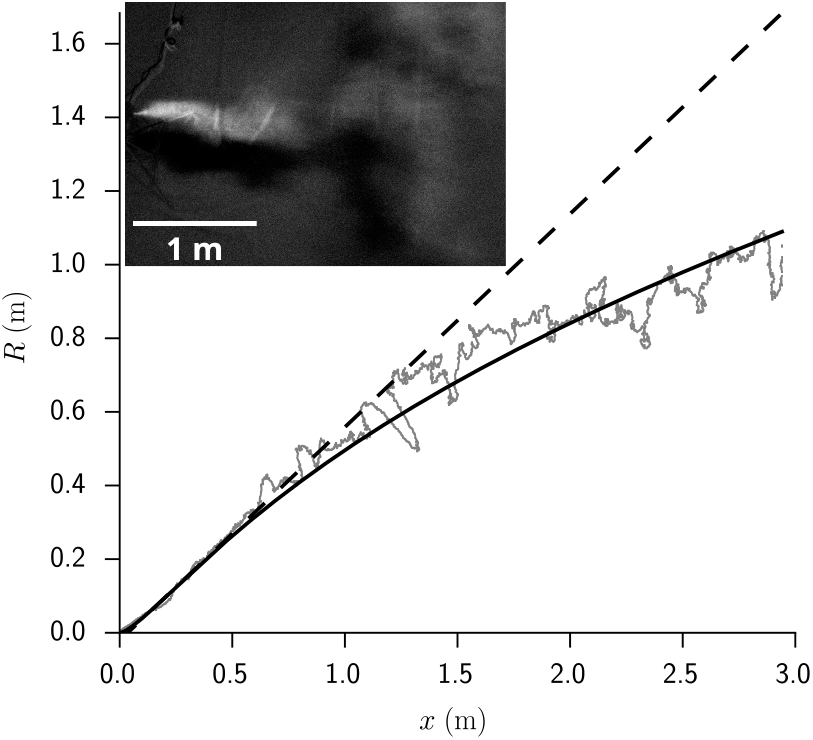
Radial extent *R*(*x*) of a turbulent fog cone, determined using video imaging from above. The contour corresponding to a scattered light intensity equal to *e*^−1^ ≈37 % of the maximum is chosen. The solid line is the solution of Eq. (25), including both the Taylor (Eq. 26) and Kolmogorov (Eq. 28) dispersion regimes. The dashed line is the tangent at the inflection point. Insert: picture from the top, showing the light intensity diffused by the fog, averaged over 0.2 s.

After an initial short transient time, a concentration field in a statistically steady state is established. CO_2_ concentration in excess has been measured in eight different locations, in corridors, near shops and a food court. The draught wind velocity *ū* was measured using a hot-wire anemometer. It ranges from 0.1 to 2 m*/*s depending on the location in the shopping malls. Measurements were done with different ventilation flow rates *Q* and recycled air fractions; the use of fire safety ventilation, namely mechanical smoke extractors and smoke vents, has been tested when available. Measurements have been performed with entrance doors both open and closed, as open doors create large draughts in some locations. Control sensors (not shown in figure 5), placed immediately upstream, to the left and to the right of the source showed no concentration increase: convection dominates over diffusion.

### 5.2 Results

Concentration profiles are averaged over the fraction of the time during which the “wind” is reasonably aligned with the sensor axis. They show a fast decay whose best fit by a power law typically gives *x*^−2^. Denoting by *R*(*x*) the typical radius of the contaminant dispersion cone (Fig. 6), the mass conservation equation provides the scaling law:

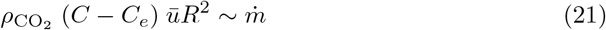

where 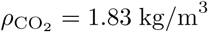 is the density of sublimated CO_2_. The decay of *C* − *C*_*e*_ is therefore consistent with the overall conical shape of the dispersion zone: the dispersion radius *R*(*x*) is roughly linear in *x*. This must not be confused with the conical shape of high speed jets in a fluid at rest. Here, the CO_2_ is dispersed by a statistically homogeneous turbulent flow.

In figure 7, the CO_2_ concentration is rescaled by 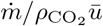 and plotted as a function of space *x*. The reasonable collapse between data shows that the CO_2_ concentration is proportional to the inverse wind velocity *ū*. The fraction of fresh air injected in the ventilation system, which controls the long-range risk, had no effect on the measured concentrations: only the local airflow disperses CO_2_. Our first conclusion is therefore very simple and directly useful for risk reduction of large public spaces: ventilation controls the long range transmission risk and fire safety ventilation is a useful way of reducing it; horizontal draughts are the predominant airflow that controls turbulent dispersion, and not mechanical ventilation.

**Fig 7.**
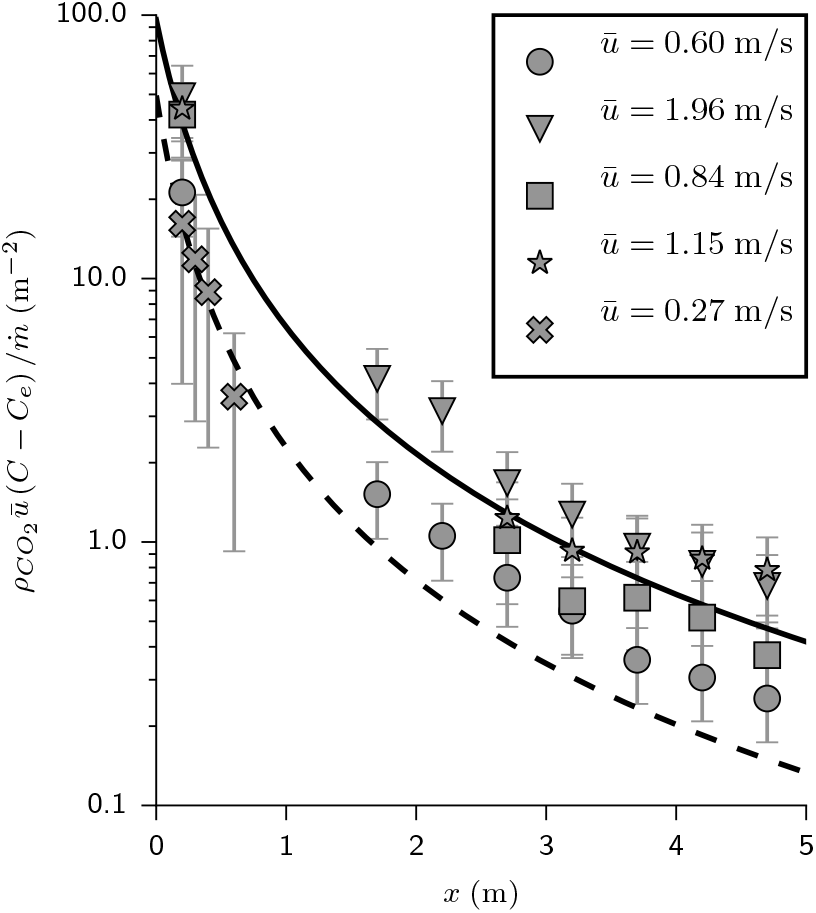
Time-averaged concentration excess profiles as a function of distance *x* to the source. The concentration is rescaled by 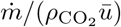, where 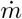 is the mass injection rate and *ū* the air velocity. The solid curve is the best fit by equation (22). Crosses and dashed line: human breathing measurements 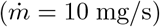 and its best fit curve.

**Fig 8.**
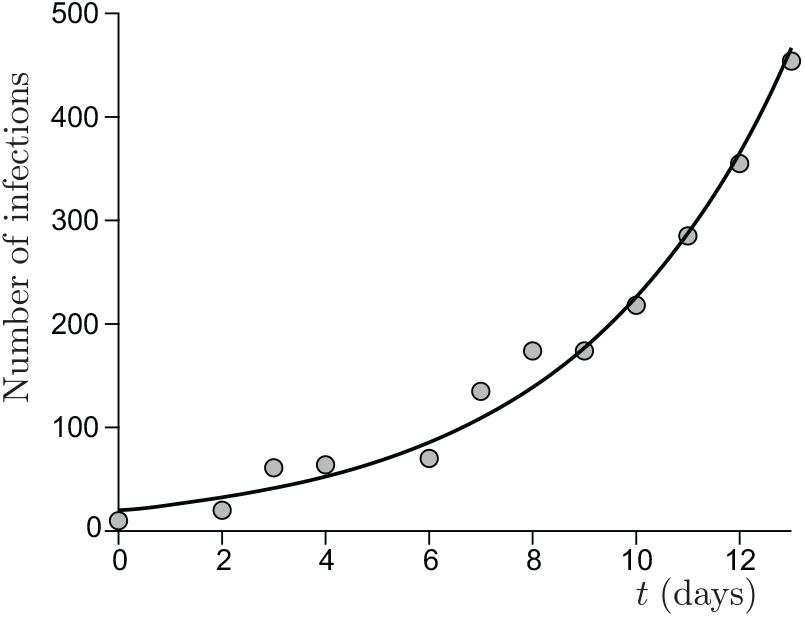
Curve relating the number of infected people onboard the Diamond Princess Cruise Ship as a function of time. The evolution is exponential (solid line), with a growth rate *σ* = 0.24 day^−1^ that corresponds to *R* = 3.9 [20, 51, 52].

**Fig 9.**
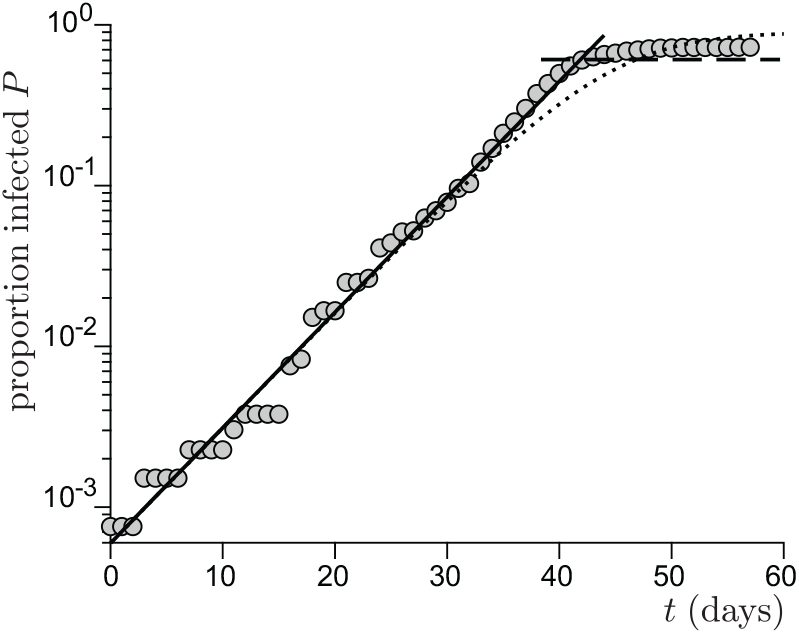
Curve relating the proportion *P* of infected people onboard the French aircraft carrier Charles de Gaulle, as a function of time *t* [53]. The solid line is the best fit by an exponential growth, which gives a growth rate *σ* = 0.16 day^−1^ that corresponds to *R* = 2.6. The horizontal dashed line is the theoretical herd immunity limit *P* → 1 − 1*/R*. The dotted line is the numerical integration of equation 19.

It has been argued on a phenomenological base by Villermaux et al. [47–49] that the turbulent dispersion cone can be geometrically described by the scaling law: *R*(*x*) ∼ *a* + *x*, at least when there is a typical separation of scale of 1 : 10 between the source size and the integral length. *a* is the effective aerodynamic source size. We can therefore parametrize the dilution by:

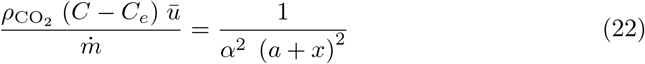

where *α* is the dispersion cone slope, determined by the turbulent fluctuation rate *u*_∗_*/ū*.

We find, *a* = 0.35 m, which is close to the actual diameter of the dry ice container and *α* = 0.10. We have also included in Fig. 7, the CO_2_ concentration in the wake of a volunteer breathing through the mouth, in a fan-induced wind of velocity *ū* = 0.3 m*/*s. We find *a* ≈0.27 m and a slightly higher fluctuation rate *α* = 0.14. The safety distance concept (2 m in French guidelines as of April 2021) does not take into account the dispersion rate, controlled by the typical flow velocity. For the gentle draught of shopping mall corridors *ū* ≃ 0.2 m*/*s, the supplementary risk at this distance corresponds to a carbon dioxide concentration excess of 50 ppm.

### 5.3 A model for short-range transmission

The scaling law *R*(*x*) ∼ *a* + *x* is a very striking approximation. Indeed, it is known since an article by Taylor [50] published one century ago that turbulent dispersion is diffusive. Any contaminant like CO_2_ or light viral particles move with the local velocity, which includes fluctuations about *ū*. Their overall motion is a random walk induced by turbulent fluctuations superimposed to a drift downwind at the velocity *ū*. The height above the ground controls the correlation length of velocity fluctuations, as this geometrical distance restricts the size of the turbulent eddies. The motion of viral particles (or CO_2_ molecules) is a Brownian motion at scale larger than the correlation length *L*. The eddy diffusivity *D* is then the product of *L* by the shear velocity *u*_∗_, which reflects the turbulent fluctuation intensity. As the Von Kármán constant is almost equal to the turbulent Schmidt number, the multiplicative factor is close to 1.

Neglecting the longitudinal diffusion, the average concentration *C* obeys the convection-diffusion equation

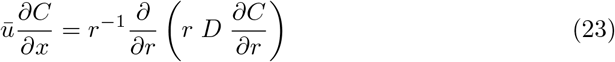

Assuming that the turbulent diffusion coefficient *D* does not depend on *r*, the equation admits an exact solution:

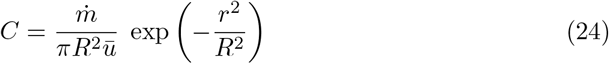

where *R* obeys the equation:

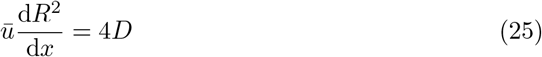

It integrates into *R*^2^ = 4(*D/ū*)*x* in the regime described by Taylor, or equivalently:

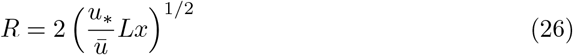

The concentration therefore decays as 1*/x* and not 1*/x*^2^ as observed.

The deviation to Taylor diffusion theory comes from the fact that the source diameter is in the inertial subrange of turbulence [47–49]. Following Kolmogorov scaling law, the contaminant is dispersed by eddies of typical size *R*, with a typical velocity difference increasing as *R*^1*/*3^. In this inertial regime, one therefore expects a scaling of the form:

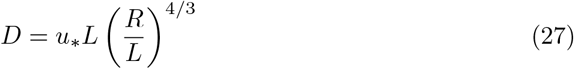

which integrates into:

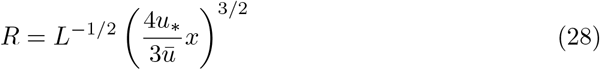

As the separation of scales between the source size and the integral length of turbulence *L* is small, the dispersion takes place in a cross-over regime between Taylor and Kolmogorov scaling laws. As equations (26) and (28) do not present the same concavity, the curve extrapolating between the two regimes presents an inflection point around which the linear approximation 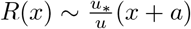 is excellent. Figure 6 shows the light intensity scattered by a fog composed of water and glycerol droplets. The dispersion in an artificially ventilated corridor is imaged from above. The experimental profile *R*(*x*) clearly shows the deviation from a quasi-conical shape at short distance towards a diffusive regime. It is compared with the solution of Eq. (25), using a phenomenological cross-over between Taylor and Kolmogorov regimes: technically *R*^−1^ is approximated as the sum of the inverse of equations (26) and (28). The approximation by a tangent at the inflection point, which corresponds to a diffusion coefficient *D* ∼ *u*_∗_*R* and *α* ∝ *u*_∗_/*ū* provides a sufficient approximation for the problem tackled here.

### 5.4 Turbulent mixing in a small room

In the previous paragraphs we have addressed the generic case in large public spaces where the dispersion is created by horizontal draughts. For completeness, we provide here a simple framework to determine the short-range risk in smaller rooms. We consider the opposite limit where there is no mean flow at all, but convective plumes creating turbulent mixing. For simplicity, we can assume that dispersion is isotropic and write the diffusion equation in spherical coordinates:

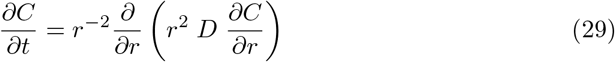

Again, considering a constant source of mass rate 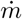, a steady state solution gradually appears, which obey:

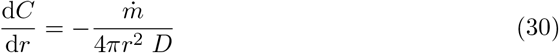

At large scale, *D* = *u*_∗_*L* can be considered as constant so that *C* decreases as *r*^−1^. In the intermediate range of scales, using the Villermaux approximation *D* = *u*_∗_*r, C* decreases as *r*^−2^. The scaling laws derived before therefore still holds, but with a geometrically determined constant *α*.

## 6 Quanta generation rate and dose response function

The risk assessment presents an interest if the quanta generation rate is known with an accuracy comparable to that on the epidemic reproduction rate *R*, say 10 %. The quanta generation rate is an average over individual properties. We must therefore examine the possibility to measure this key quantity, statistical by nature, starting from epidemiologic and from virologic measurements.

### 6.1 Epidemic growth rate in a microcosm

The fast contamination in the Diamond Princess boat [20] has provided the first proof of airborne contamination by SARS-CoV-2. It has been deduced from the cold weather conditions (−5 ^°^C) that the ventilation was mostly recycling the air, at 70%. Even with 30% of fresh air, this would be an excellent ventilation rate if the air conditioning had been equipped with HEPA filters removing most of particles in the 100 nm −1 *µ*m range. The persistence of viral particles in such circumstances is limited by their deactivation timescale, around 1 hour. The surface accessible to passengers is 78 10^3^ m^2^ and the ceiling height is 2.4 m. The indoor volume, 187 10^3^ m^3^, is pretty large compared to the number of people, *N* = 3711, crew and passengers together. The epidemic growth rate was *σ* = 0.24 day^−1^ over 12 days. According to equation (20), the epidemic reproduction rate is therefore *R* = 3.9.

From equation (18), this corresponds to a total viral emission 𝒩= 400 quanta or, equivalently, to a typical viral emission rate 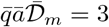 quanta*/*hour. Considering the viral load curve *h*(*t*), this value corresponds to an emission rate on the order of 16 quanta*/*hour at maximum.

The fast contamination in the Charles de Gaulle French aircraft carrier is similarly due to the lack of filtration of the recycled air. Over 1767 people onboard, crew and commandos together, 1288 were infected in a short period of time. As for the Diamond Princess, the persistence of viral particles is limited by their deactivation timescale rather than by the ventilation. The indoor relevant volume is estimated around 150 10^3^ m^3^. The epidemic growth rate was on the average *σ* = 0.16 day^−1^ during the epidemic peak. According to equation (20), the epidemic reproduction rate is therefore *R* = 2.6.

From equation (18), this corresponds to a total viral emission 𝒩= 460 quanta or, equivalently, to a typical viral emission rate 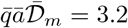 quanta*/*hour. Considering the viral load curve *h*(*t*), this value corresponds to an emission rate on the order of 18 quanta*/*hour at maximum.

### 6.2 Contribution of schools to the reproduction rate

Schools constitute the best documented social sub-system [20]. Contrarily to the previous case of boats, schools are not isolated from the society. United Kingdom provides the best data sets for both systematic PCR tests and ventilation conditions. Figure 10 shows the epidemic evolution for secondary school pupils, between lockdown and holidays. It allows one to estimate the contribution of schools to the epidemic rate, in the absence of mandatory masks, to *R* = 1.7 in March 2021. CO_2_ concentration have been measured in British schools during a year, showing an average of *C* = 1070 ppm in March. Figure 11 shows the typical evolution of this concentration in a classroom whose volume is 10 m^3^ per pupil. Considering that the average school time for secondary schools is 27.5 hours per week, equation (18) allows one to deduce the total viral emission 𝒩= 230 quanta. It corresponds to a typical viral emission rate 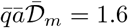 quanta*/*hour and an emission rate at maximum on the order of 9 quanta*/*hour.

**Fig 10.**
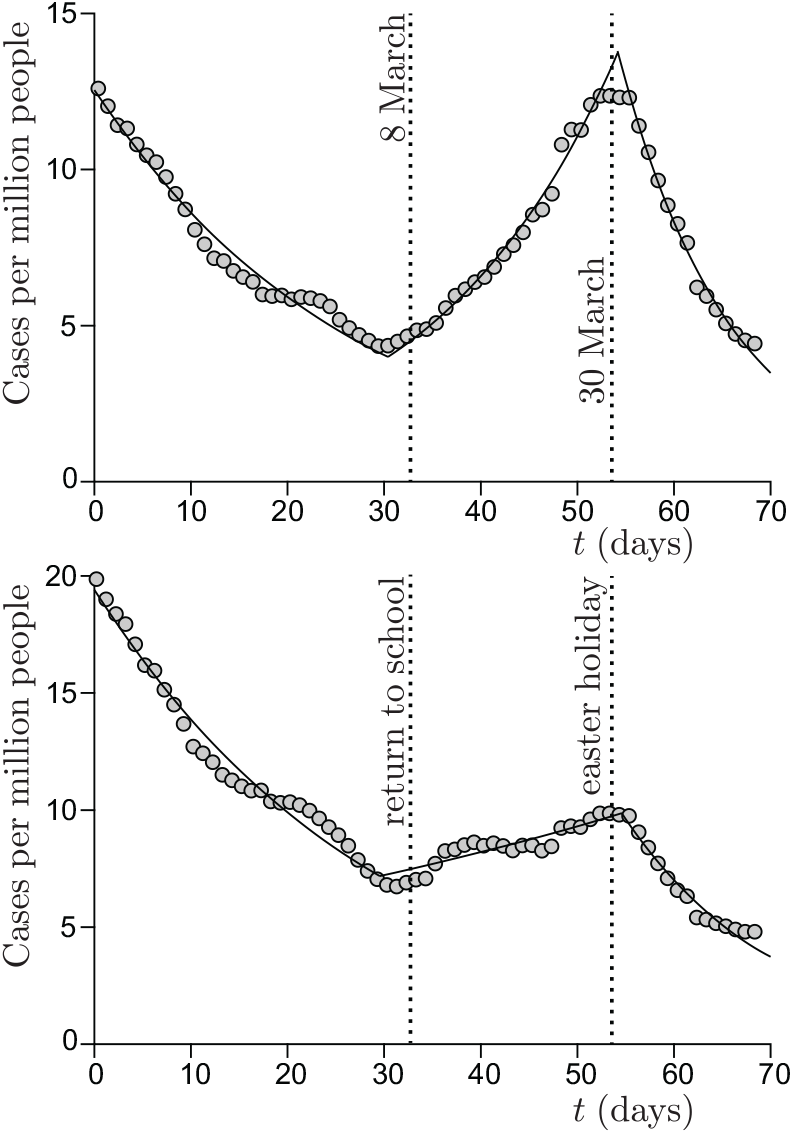
Cases per million people in United Kingdom, from 1 February to 12 April. (a) Pupils from age 10 to age 14, with no mandatory mask. The best fit by an exponential provides the reproduction number: *R* = 1.34 ± 0.04 during school period vs *R* = 0.81 ± 0.03 before and *R* = 0.70 ± 0.03 after. (b) Pupils from age 15 to age 19, with mandatory masks. The best fit by an exponential provides the reproduction number: *R* = 1.07 ± 0.04 during school period vs *R* = 0.82 ± 0.03 before and *R* = 0.70 ± 0.03 after.

**Fig 11.**
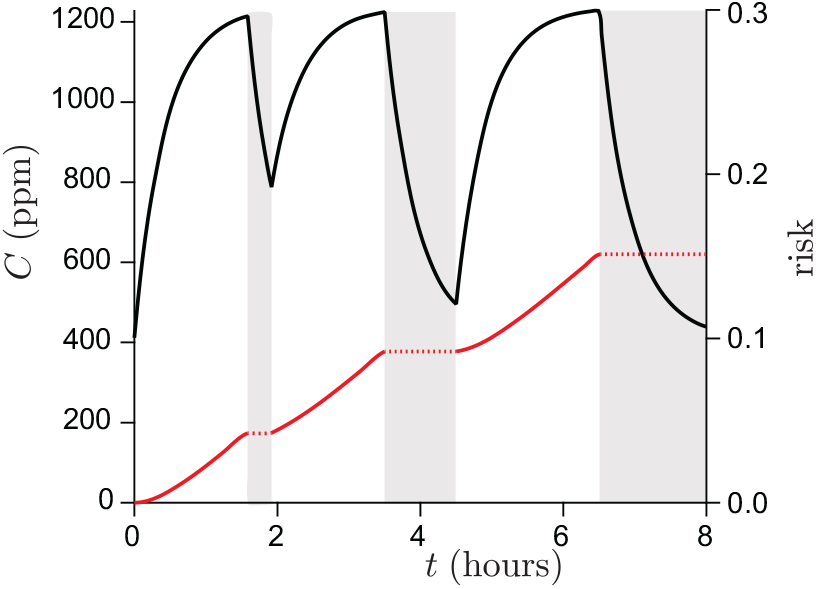
Typical ventilation in British schools in March, as deduced from Vouriot et al. [54]. Left axis: CO_2_ concentration as a function of time. The pupils are not present in the classroom during the periods of time shown in gray. Right axis: deduced risk *r*.

**Fig 12.**
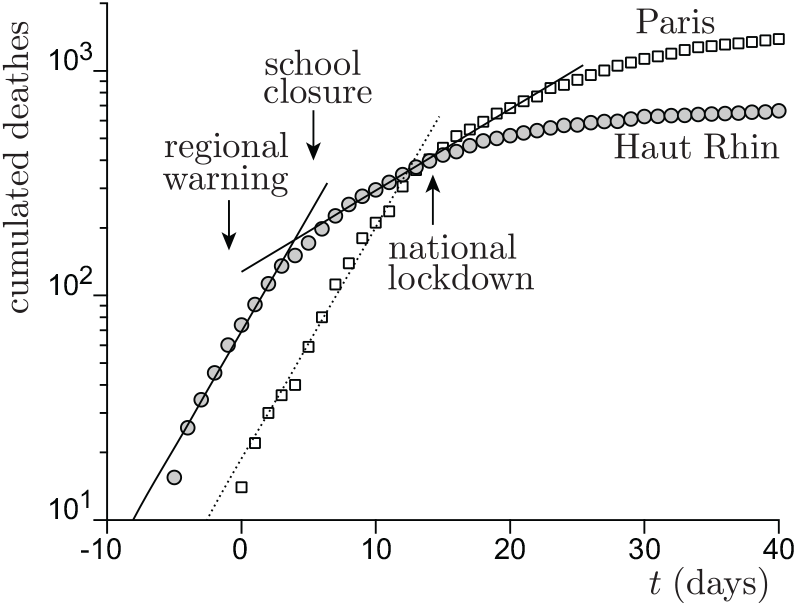
Cumulated number of deaths due to Covid-19 as a function of time *t* in two French departments, Paris (white squares) and Haut-Rhin (grey circles), in March 2020. The delay between lockdown and the maximum of death rate was 17 days. Arrows show the effect of school closure in Haut-Rhin, one week before national lockdown of March 17, lagged by this delay. During the week when school closure has been the single measure taken against the epidemy, the reproduction number has changed from *R* = 3.86 ± 0.1 to *R* = 1.61 ± 0.1.

Importantly, the contribution of school to contamination is half smaller (*R* = 0.39) for pupils from age 15 to 19, while the epidemic rate is the same during lockdown and holidays. We ascribe this reduced risk to mandatory masks, as discussed later.

In France, in February 2020, the SARS-CoV-2 epidemic emerged first in the Haut Rhin department (Alsace) and a few days later in Paris. The alert was given in local newspapers on March 6. During one week, before the national lockdown on March 17, schools have been closed in Haut Rhin and no other health risk reduction measure was taken. The overall effect of this single measure has been the reduction of the number of cases by 60%. Indeed, in Paris, no effect is observed before the national lockdown. The overall effect of school closure on the epidemic rate, Δ*R ≃* −2.25, shows that each contamination of a pupil at school, for an ordinary social live, leads to 0.7 secondary infections inside school and 1.55 outside it.

### 6.3 Super-spreading events

The epidemic outbreak in a poorly ventilated restaurant (0.7 ACH) of Guangzhou [13, 55] has been entirely characterized from the aerodynamic point of view. Air conditioning was producing a contaminated recirculation bubble isolating a subpart of the restaurant with *N* = 26 customers. Both the dilution factor *ϵ* and the duration of inhalation of contaminated air are known for all customers. Taking into account the accumulation of viral particles over time, we can find the emitted dose per unit time for which the number people that are contamined on the average is exactly that observed: *M* = 9. We find a viral emission rate of 40 quanta*/*hour.

The Ningbo Tour Bus is an example of super-spreading event with null ventilation [20]. 20 people have likely been infected amongst *N* = 68 in the bus going to and coming back from a religious ceremony in *t* = 50 minutes. The bus volume was *V ≃* 45 m^3^. Inverting equation (8), the average dose inhaled by all bus passengers is *d ≃* 0.35 quanta. The viral emission rate is therefore 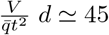 quanta*/*hour.

In both cases, the viral emission rate for these super-spreading events is 2 to 3 times larger than average, which sounds reasonably consistent.

### 6.4 Molecular determination of the infectious quantum

The viral dose can be expressed in RNA molecules (GU, for viral genome units) using quantitative RT-PCR. The result of RT-qPCR is expressed in number of threshold cycles (Ct) which is transformed into GU (or copy of viral RNA molecule) after calibration using a solution of viral RNA molecule of known concentration. Externally, in a drop, the number of RNA molecules is probably a good proxy for the number of virions (one genome per virus particle), but not all of them are liable to produce an infection. In order to express a infectious quantum in GU, it is necessary to measure a dose-response curve relating the fraction of contamination of a population to the inhaled dose of RNA molecules, in GU. The Wells-Riley model is a popular yet uncontrolled approximation of the equation (7) governing the dose-response curve in the absence of viral cooperativity: given the linearization (8) and the limit at large dose 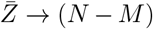, it assumes that the dose response function remains exponential:

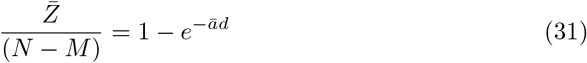

where *ā*^−1^ is the infectious quantum. However, the dose response curve may tend to 1 much more gradually in a population presenting a wide distribution of immune response. The ID50 is defined as the infectious dose necessary to get a 50% contamination. In the Wells-Riley model, the ID50 is ln(2) ≃ 0.69 quanta.

The dose-response curve depends on the characteristics of the virus, the route of inhalation and the organism studied. It is well understood that there is little or no data on human populations, especially when the virus presents a risk of lethality. To approach this curve, organisms close to humans (non-human primates) and viruses of the same family (the coronavirus family for SARS-CoV-2) may be considered, assuming that the dose response curve is mainly determined by the cellular entry point. A complementary approach is to measure the contamination of a cell monolayer by a viral solution in which the concentration in RNA molecules is known. One counts the number of plaque-forming units (PFU), which corresponds to the number of virions from the initial inoculum that will lead to a hole in a cell monolayer, by killing their host cell, multiplying, disseminating to neighbouring cells and destroying them in turn. This number, in PFU, can be used to express the viral titre in a solution or in a drop, or to express an inhaled dose. Here, only the virulent viral particles, which are effective for infection, are numbered, in contrast to GUs which encompassed both infectious and non-infectious particles (for instance, damaged or dried-out virions). The ratio between plaque-forming unit (PFU) and RNA molecules (GU) in a given sample, varies from study to study, depending on the nature of the inoculum tested, on the quality of its preparation and preservation, and on the infection conditions. Alternatively, the ability of the virus to kill cells can be measured by relating the viral activity to the viral dose expressed in GU. This curve is usually characterized by the Tissue Culture Infectivity Dose giving 50% of the response (TCID50), also referred to as the viral activity. Again, one expects the relation 1 TCID50 = ln(2) PFU, any significant deviation indicating a protocol problem. For SARS-CoV-1, one plaque-forming unit (PFU) ranged between 1200 and 1600 GU [56]. For SARS-CoV-2, the literature provides two measurements of the plaque-forming unit equal to [57, 58] 1000 GU and 1500 respectively for samples of high preservation quality. We may therefore use 1 PFU ≃ 1400 GU as a reasonable ratio during the asymptomatic phase of the pathology. Upon the course of the disease, the number of replicable virion is going to decrease and this ratio will increased.

The ratio between ID50 and TCID50 is larger than 1 and relates the viral dose able to infect an organism to that able to kill a culture cell. To infect an organism, the coronavirus must infect the cells of the epithelium, replicate and not be eliminated by the immune system of the upper respiratory tract, the mucus. For SARS-CoV-2, this ratio ranges from 10 to 1000 depending on individuals. [59] We are interested here in the average over the population, which presents large error bars around the best estimate determined on SARS-CoV-1 that may be used [60], ≃ 350. In conclusion, the infectious quantum, defined over a population is around *ā*^−1^ = 1 quantum ≃ 5 10^5^ GU within, say, a factor of 2.

In order to express the exhaled viral dose per unit time during breathing in quanta*/*h, the quantity of SARS-CoV-2 RNA per unit air volume exhaled by patients must be measured. In the study by Ma et al. [61], patients were asked to exhale toward a cooled hydrophobic film via a long straw to collect a sample of exhaled breath condensate: the SARS-CoV-2 concentration was in the range 10^5^ −2 10^7^ GU*/*m^3^. As a consequence, the viral emission rate of SARS-CoV-2 in the breath ranges from 0.1 to 20 quanta/h. This value relies heavily on the ratio between ID50 and TCID50 that we have used. In the same article [61], the authors notice that the emission rate was correlated with the viral load in the nose and the throat but not in the lung. The viral exhalation rate varies in time and reaches its maximum during the asymptomatic/presymptomatic phases.

## 7 Respiratory mask efficiency

### 7.1 Droplet size distribution

Coughing, sneezing, singing, speaking, laughing or breathing produce droplets of mucosalivary fluid in two range of sizes. Droplets above 100 *µm* are produced by fragmentation of a liquid sheet formed at the upper end of the respiratory tract (i.e. for sneeze) or from filaments between the lips (i.e. plosives consonants when speaking [62, 63]), with an average around 500 *µm* [64–66]. In the first case, the initial sheet is stretched and get pierced. The liquid accumulates by capillarity retraction in a rim, which destabilizes into ligaments [67, 68]. The latter form droplets by a capillary instability referred to as the beads-on-a-string [69]. In the second case, a film forms between the lips, which destabilizes into filaments, themselves exhibiting the beads-on-a-string instability.

Droplets below 20 *µm* form either by bubble bursting events in the lungs alveoli or by turbulent destabilization of liquid films covering the lower and upper airways. The average droplet size, around 4 *µm* results from an interplay between the fluid film thickness, the turbulent stress and the surface tension. Further fragmentation of these droplets can occur in the tract constrictions where air flows at large velocity.

Pulmonary surfactant helps reducing the droplet size, and contributes to prevent accumulation of fluid in airways. The main entry zone is through the nasal epithelium and more specifically a subset of cells of the nasal epithelium expressing both the ACE2 receptor and the TMPRSS2 protease. Other entry zones exist as well as different receptors and proteases [28]. In first approximation, they can be considered as minor routes in the dissemination of the epidemic and in the risk of contamination. The emission of mucus droplets containing viral particles in the nasal cavity has not been much investigated so far.

Between 20 *µm* and 100 *µm*, almost no droplets form, indicating two well separated mechanisms. For each class of droplets, the distribution around the average has been fitted either by a log-normal distribution, following the idea of a break-up cascade, or by a Gamma distribution, based on the idea that the blobs that make up a ligament exhibit an aggregation process before breaking up [70].

### 7.2 Evaporation process

The evaporation of liquid droplets in the air is controlled by the ambient relative humidity RH. Mass transport of water molecules from the droplets to the surrounding air is diffusive; as the drops evaporate, they release latent evaporation heat, which is also conducted away. This cools down the droplets, which in turns lowers the saturation pressure in the immediate surrounding of the drop, slowing down evaporation. Due to this coupled transport, a drop of initial radius *a*_0_ shrinks to a radius *a*(*t*) as [71, 72]

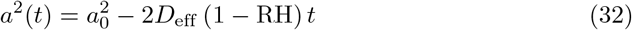

where *D*_eff_ = 1.3 10^−10^ m^2^*/*s is an effective diffusion coefficient taking into account both diffusive transport of mass and its slowdown due to evaporative cooling. Evaporation takes places at the surface of the drop, which explains the linear behaviour of *a*^2^ with *t*. Since the drops are small, evaporation is very fast compared to the time they spend aloft: a 4 *µ*m drop at 70 % RH completely evaporates in 0.2 s, while it takes 20 min to fall a distance of 2 m under its own weight.

The classical picture [73] considers evaporating droplets as independent. This is true for aerosol droplets dispersed inside a room, but not of droplets inside a cough or sneeze spray. In that case, RH is roughly uniform and close to 1 inside the aerosol jet, meaning that no evaporation takes places except at the spray boundaries [74, 75]. This makes these drops extremely long-lived, up to a hundred times the isolated drop lifetime [72, 76, 77].

However, virus-laden respiratory droplets do not vanish as they contain viral particles and are not composed of pure water. The mucosalivary fluid is a dilute solution of surfactants, proteins and electrolytes, initially composed of ∼ 99 % water in volume. The solutes stabilise drops at a finite radius *a*_eq_, at which they still contain water [78]:

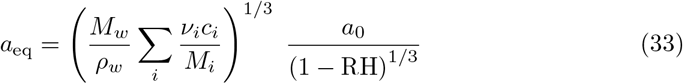

The sum is done over all solutes *i. c*_*i*_ is the mass concentration of solute *i, M*_*i*_ its molar mass, *ν*_*i*_ its degree of dissociation (2 for NaCl). We model respiratory fluid by a mixture of NaCl and the total average protein content [79, 80]: *c*_electrolytes_ = 9 g*/*L (physiological NaCl concentration), *c*_proteins_ = 70 g*/*L, *M*_proteins_ = 70 kg*/*mol. This gives *a*_eq_ ≈ 10^−1^*a*_0_ (1 − RH)^−1*/*3^: typically, at medium relative humidity, aerosol drops formed at 5 *µ*m remain at 500 nm, which is significantly larger than the virus itself.

In aerosol droplets at equilibrium, virions are gradually inactivated by the damage done by dessication or by antiviral proteins in saliva [81]. The inactivation rate of envelopped, airborne viruses increases [82–84] with RH: this suggests that virions can associate with proteins which protects them both from dessication and antivirals [80]. By stabilising the droplet at a finite radius, solute reduce the evaporation time to

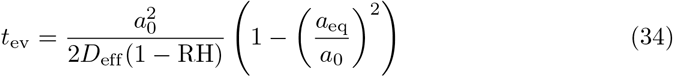

The solute effect on the evaporation time is small at low RH since the drop shrinks by a lot; at 99 % RH, a 4 *µ*m drop has its evaporation time increased by 80 %.

### 7.3 The enrichment issue

In the literature [20, 54, 85–89] devoted to airborne transmission of SARS-CoV-2, most authors have considered that the viral emission rate is the product of the volume of mucus droplets emitted per unit time by the concentration in viral RNA in mucus. The viral particle emission rate was found far too small. The volume of droplets emitted per unit time has been measured at short distance from a person breathing (in the range 20 −2000 *µ*m^3^*/*s) or speaking (∼2000 *µ*m^3^*/*s) [64, 72]. This would correspond, if the droplets were composed by mucus with the same viral charge as in a nasopharyngeal swab, to an emission of 40 − 4000 GU*/*h. This range of values is more than three orders of magnitude smaller than direct measurements of viral particles.

We propose here two possible explanations to this discrepancy. First, small droplets evaporate very quickly so that the droplet diameter may be much smaller when measured than when generated by instability. A reduction of the drop size by a factor of 10 would be sufficient to explain the underestimated mission rate. Second, during normal breathing, viral particles are emitted from the nasal mucus. They may serve as surface disturbances favoring aerosol formation encapsulating viral particles. This enrichment procedure would then lead to a viral content of droplets which is not proportional to their initial volume. If droplets of 4 *µ*m were composed of a mucus containing 10^8^ GU*/*mL, only one in 300 aerosol droplets would contain a viral RNA. The possibility that drops nucleate on viral particles cannot be rejected on the basis of a simple order of magnitude.

It is worth noting that the total quantity of liquid exhaled per volume of air is around 30 mL*/*m^3^. If all this liquid was initially under the form of droplets, then the viral emission rate would be 8 10^9^ GU*/*h, which is two orders of magnitude too large. Just like evaporation creates a bias in the droplet sampling, condensation of vapour in the drop sampler is a symmetric problem [90].

### 7.4 Masks

The filtration factor *λ* crucially depends on the size of mucus droplets carrying viral particles. All types of masks totally filter droplets of a fraction of millimeter. However, they have very different efficiencies between 0.1 *µ*m and 0.5 *µ*m, which is the range of equilibrium sizes of mucus droplets after evaporation. Mask efficiency presents a minimum around 0.3 *µ*m. Smaller particles diffuse due to brownian motion, which increases the probability of collision with a fiber. Larger particles present a higher collisional cross-section with fibers. The filtration efficiency is typically 95 % for FFP2/KN95 masks, 70 % for surgical masks and 20 % for community masks for this range of particle diameters while they all present a 100 % filtration above 10 *µm*. We have ourselves performed filtration measurements of incense smoke whose size distribution ranges from 0.1 *µ*m and 1 *µ*m. The smoke was sampled with and without mask in a transparent air sampling canister, prepared by evacuating the contents to a vacuum of approximately 100 mbars. The filtration efficiency is deduced from Beer-Lambert law, using the attenuation of a LASER whose intensity is measured with a photodiode. The effective filtration efficiency was 97 ± 2 % for FFP2/KN95 masks, 94 ± 2 % for surgical masks and 83 ± 3 % for cotton masks distributed in French universities. This is similar to the values obtained by Fischer et al. [91].

The main problem of face masks is leakage along the nose and the cheeks. A well fitted surgical mask as an aerosol filtration around 90 % which corresponds to *λ* = 0.1 but ordinary wearing is much lower (*λ ≃* 0.3 − 0.7). Community masks only protect against the largest aerosol droplets (*λ ≃* 0.7). FFP2 are much easier to fit and it is possible to reach effectively the value *λ* = 0.95 giving the name to KN95 [92]. More precisely, the pressure drop across FFP2 helps fitting the mask to the face when inhaling but favors leaks when exhaling, as shown in figure 13.

**Fig 13.**
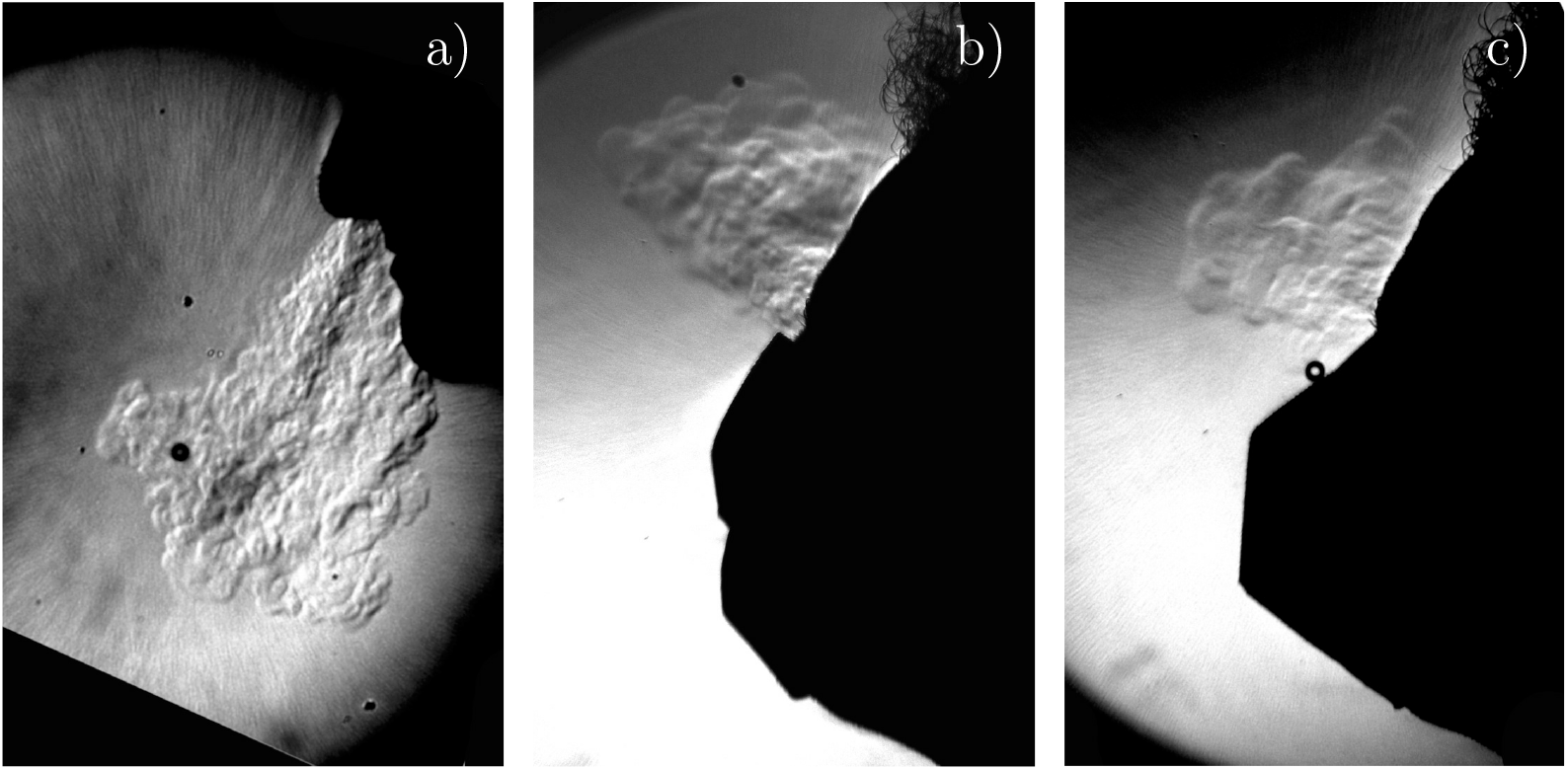
Schlieren imaging of a person exhaling (a) without a mask (b) with a surgical mask (c) with an FFP2 mask. Temperature acts as a passive scalar with respect to turbulent transport, the same way CO_2_ and small aerosols do. The schlieren technique shows local variations in the air refractive index caused by the warm air exhaled out of the body.

We have performed a quantitative study of mask wearing in different public spaces around Paris. Figure 14 shows the resulting histogram reflecting social activities at different times of the week, in April 2021. Only indoor public spaces where masks are mandatory have been included. FFP2 masks represent 3 % only of facial masks and correctly fitted protective masks (chirurgical or FFP2) only 42 %. We can estimate that the mean filtration factor *λ* is around 0.5 only, which gives a reduction of *R* by a factor of 4 in public spaces were mask is worn. The reproduction rate in France has started around *R* = 3.8 in February 2020 and ended around *R* = 0.9 with the wild strains, before the strain B.1.1.7 became dominant, one year later. Following the same reasoning as before, we may consider that each contamination outside the family and friends circle (people than one meets on a regularly basis without facial masks) is amplified by a factor 2.2. Under this hypothesis, the mean number of secondary infection outside this close circle has decreased from 1.75 to 0.4, a factor of 4.

**Fig 14.**
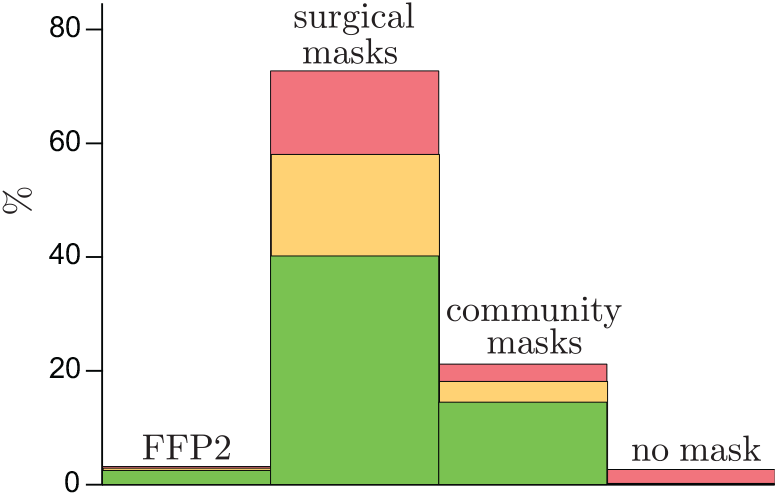
Mask wearing in five selected public spaces in the Paris suburban region (Ile-de-France) where facial masks are mandatory, mid April 2021, during lockdown (supermarkets and public transportations). *N* = 584. Green: correct fitting. Orange: centimeter-scale leakage along the nose or the cheeks. Red: No fitting at all.

Figure 10 shows that secondary infection in British schools was around 0.6 without facial masks and 0.3 with mandatory masks. We have observed that young people statistically wear their facial mask in a non fitted way. This may explain a part of this reduction of risk by a factor 2 only. An alternative explanation is the contamination during lunch time, which remained unchanged.

A study in Finland [93] has shown an infection rate of healthcare workers substantially higher (more than 15 times) than that of the general population. Amongst 413 healthcare workers who performed aerosol-generating procedures with Covid patients during spring 2020, 14 occupational infections occurred using surgical masks (amongst 233) and 0 with FFP2/FFP3 masks (amongst 180). This suggests a higher efficiency ratio between FFP2 and surgical mask than expected, due to leakages.

## 8 Conclusion

### 8.1 Summary of results

In this article, we provide an effective definition of the risk *r* associated with a public space, defined as the average secondary infections per initially infected person. Under the commonly accepted hypothesis of no cooperation between virions, the risk is computed in the low risk limit. It is related to the integrated quantum emission 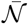, to the mask filtration factor *λ* and to the CO_2_ concentration, which quantifies the dilution factor between exhaled and inhaled air. The first central result of the article, given by equation (14), is the incorporation of the ventilation flow rate, the room volume and the number of people present into a single measurable quantity. The disappearance of the number of people *N* from equation (14) at large *N* results is non-trivial and comes from two factors balancing each others: on the one hand, CO_2_ is exhaled by all individuals present, and not only people infected by the virus; on the other hand, the transmission risk increases linearly with the number of people susceptible to be infected.

Indoor and outdoor spaces both present a risk of airborne transmission at short-range, in the dilution cone of the exhaled breath. The concentration in viral particles or in CO_2_ decays as the inverse squared distance to the emitter, and as the inverse wind speed. Figure 15 summarizes the findings: the risk, higher in the wake of other people, at short distance, can be determined by adding a short range excess CO_2_ concentration to the well-mixed case.

**Fig 15.**
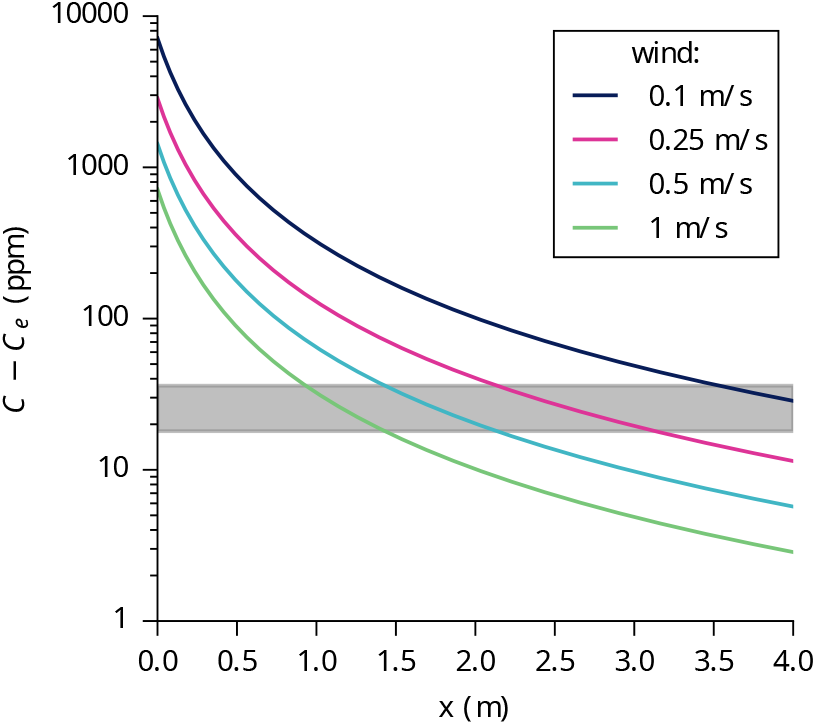
Modeled CO_2_ concentration excess in the wake of a person as a function to the distance *x* in the wind direction, for different wind speeds *ū. ū* = 0.1 m*/*s is the smallest natural wind found inside shopping mall corridors; 0.5 m*/*s is a moderately windy corridor; 1 m*/*s is light air outdoor. Gray rectangle: concentration range such that the risk obey 0.5 *< r <* 1 without facial masks. The security distance, given by the intersection between gray band and colored lines strongly depends on the flow velocity.

The viral load curve results into an important source of variability. Here, we have defined the integrated, maximal and average viral emission rate, which are significantly smaller than previous estimates: 3 quanta*/*hour for the raw strain, which corresponds to 16 quanta*/*hour at maximum and 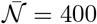 quanta for the integrated quantum emission. These values must be multiplied by ∼1.5 for the B.1.1.7 strain and by ∼2.0 for the P.1 and B.1.617 strains [46]. The mean viral emission rate is consistent with both epidemic and molecular measurements. Importantly, the infection dose is around 5 10^5^ GU and not between 10 and 100 as mentioned in some recent articles [20, 26, 54, 85–89], once the ratio between plaque-forming units (PFU) and genome units (GU) taken into account.

### 8.2 How to reduce the epidemic risk in public spaces?

The transmission of SARS-CoV-2 in public spaces is predominantly airborne. The infection risk can be reduced by a combination of five actions:

- Correct wearing of good quality masks to increase the mean filtration factor *λ*.
- Ventilation with a sufficient fresh air flow rate per person to reduce the long-range risk.
- Air purification to complement ventilation where needed.
- Turbulent dispersion, distancing and reduction of static crowds to reduce the short range risk, both indoors and outdoors.
- Monitor CO_2_ to measure the risk and adjust practices.

The risk of fomite transmission after droplet deposition on surfaces is probably negligible but it is still interesting to mitigate it to prevent contaminations by other illnesses. It can be eliminated by regular hand and surface washing.

The results in this article can be applied directly to the problem of comparing risk scenarios in public spaces, encouraging rational public policies to reduce the transmission risk in such places. The uncertainty lies primarily with the mask filtration factor *λ* and with the integrated quantum emission 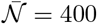 quanta, but the relative risk can be precisely determined. Still, it is interesting to compute the maximal CO_2_ concentration consistent with the receding regime (*R <* 1), following the Zero-Covid strategy. This should be considered as an initial estimate, which must be updated when better data become available. Allowing for a small safety factor, we can take 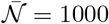 quanta for the strains that are about to become dominant in the second half of 2021 and a risk *r* = 0.5. In the absence of facial masks, the excess CO_2_ concentration must be limited to 18 ppm and therefore to *C* = 430 ppm. This is almost impossible indoors without opening all windows. Having this value in mind, figure 15 shows that the transmission risk outdoor, without masks, is real at short distances and for long periods of time. Static crowds without masks must therefore be avoided. It is important for people to learn how to take into account the wind strength and direction for static outdoors activities, in particular if they sing, eat, or drink for a long duration. As the risk outdoors is entirely at short range, large fans may be used to reduce the risk of a bar terrace or static queue in front of a shop (figure 16 panel a). The more these fans induce turbulent fluctuations, rather than an average flow, the better they are. They must be oriented upwards to change the wake direction. For outdoor dance floors, injection of air at high flow rate, say, 1 −10 m^3^*/*hour*/*person may be sufficient to reduce the risk (Fig. 16 panel c).

**Fig 16.**
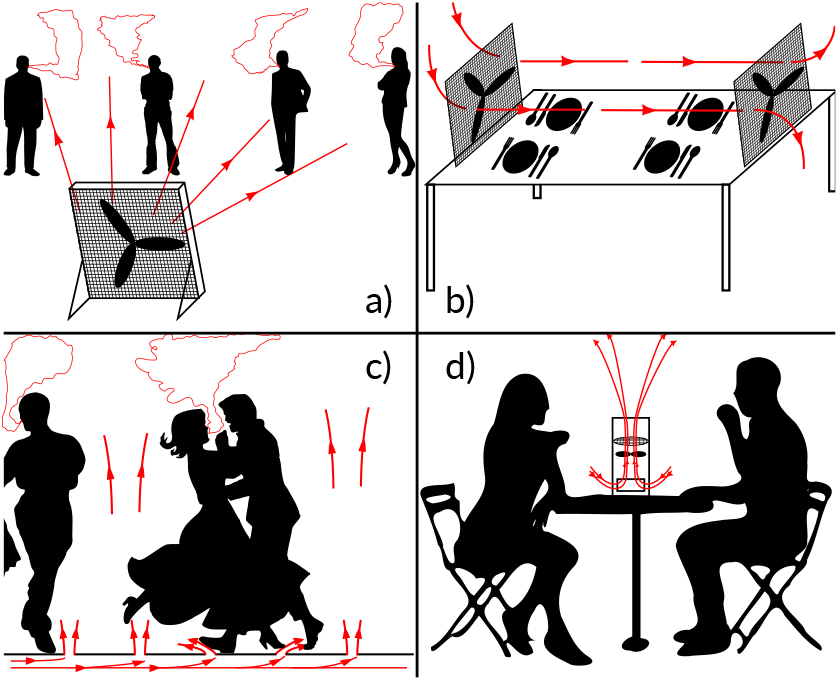
Hydrodynamic solutions to mitigate short-range transmission by dispersing aerosols away from individuals. (a) A large fan with a filter is slightly tilted upward and aimed at a line of static people waiting. The cone of aerosols they emit cannot reach anyone in the line. (b) Fans force the circulation of air through Hepa filters at a cafeteria table. (c) An air flow inside the floor of a club pushes air up through small holes, preventing aerosols from spreading laterally and dispersing them towards the ceiling where the ventilation system can remove them. (d) A fan at a café table pulls air, filters it and expels it upwards.

With the current level of mask-wearing in public (Fig. 14), which leads to a small filtration factor *λ*^2^ = 0.2, the maximum excess CO_2_ concentration should be 90 ppm, which corresponds to an unrealistic maximum of concentration *C* = 500 ppm. The reduction of risk in public spaces must therefore combine improvement of mask fitting and ventilation. For instance, with an objective of 10 % FFP2, 70 % well-fitted surgical masks, allowing for 20 % community masks or badly fitting masks, one can easily decrease the filtration parameter *λ* by a factor of 2. Explanatory signs, scientific video clips on aerosol contamination, on the correct wearing of masks and on the filtration levels of the different types of masks may encourage public adherance. For an average filtration of *λ*^2^ = 0.05, a maximal excess CO_2_ concentration of 390 ppm becomes sufficient, which corresponds to *C* = 800 ppm. This constitutes a reasonable compromise between masks and ventilation.

Consider now public places like theaters or concert halls where FFP2 masks can be included in the ticket price with different sizes available. The factor *λ*^2^ would be decreased by 40. The risk becomes negligible for standard ventilation conditions. Surgical masks and FFP2 masks are expensive but can be decontaminated about four times and reused as long as the filtration layers do not show tears. Public spaces could offer a mask decontamination service using a combination of UV-C, heat and hydrogen peroxide vapor [94].

Ventilation with fresh air is energy consuming, both in winter and during heat waves. The filtering of viral particles in the recycled air is difficult to implement because the standard ventilation installations have not been dimensioned to receive Hepa filters: they are not able to overcome the induced pressure drop. Alternatively, two innovative methods can be used and even combined to decontaminate the air: UV-C neon lights, which are already used reliably and regularly [95–103], and ultrasounds between 25 and 100 MHz [104], which is at the proof-of-concept stage. These techniques are cost-effective in the long term for destroying nucleic acids, DNA or RNA from bacteria, viruses or other micro-organisms present in the air.

The absence of masks when eating and drinking poses a specific problem of aerosol risk reduction. In particular, collective catering facilities are amongst the most important places of high transmission risk. It is possible to use Hepa filtered air purifiers arranged to provide air free of viral particles and suck out stale air (Fig. 16 b-d).

### 8.3 Concluding scientific remarks

In conclusion, it is important to discuss the limitations of this study. A better molecular determination of the infectious quantum necessitates measuring the quantity of viral particles per unit volume of exhaled air. This calls for the design and the calibration of facial masks allowing patients to breathe normally and to collect all viral particles in a filter. We have hypothesized here that the infectious quantum, expressed in viral RNA (GU), remains the same for SARS-CoV-2 variants, which differ by the viral load curve. The systematic use of breath aerosol samplers [105] would provide a quantitative characterization of SARS-CoV-2 strains, necessary for risk assessment and subsequent risk-reduction policies. Similarly, one could directly use standard quantitative techniques of molecular biology such as plaque assay and quantitative polymerase chain reaction to directly measure facial mask efficiency.

A second limit of the study is the lack of knowledge on the generation of virus-laden aerosols in the upper tract, particularly in the nasal cavity. We have shown that the indirect determination of the rate at which viral particles are exhaled using the aerosol droplets emission rate underestimates the result by three orders of magnitude.

Finally, the evolution upon desiccation of mucus droplets carrying virions and the mechanisms of deactivation of SARS-CoV-2 remain poorly understood. Evaluating the efficacy of other techniques to reduce risk requires knowing how environmental conditions (temperature, humidity, chemical concentrations, ultraviolet irradiation) affect the viability of SARS-CoV-2 [72, 77, 106–109].

## Data Availability

No external dataset

## acknowledgements

This article involves seven undergraduate students who have worked on the problem in the context of their final-year experimental physics courses (“Phy Exp”) at the Université de Paris. The authors thank the “Phy Exp” team and in particular the lathe-mill operator, Wladimir Toutain, and the technician, Thibaut Fraval De Coatparquet, for their assistance. Jérome Jovet has helped to obtain the Schlieren imaging. B.A. and J.H. thank Alice Lebreton-Mansuy and Joël Pothier for fruitful discussions. Unibail-Rodamco-Westfield has funded this work under the CNRS contract 217977 and provided access to Forum des Halles and Carré Sénart as well as technical assistance.

